# Plasma proteomics of SARS-CoV-2 infection and severity reveals impact on Alzheimer and coronary disease pathways

**DOI:** 10.1101/2022.07.25.22278025

**Authors:** Lihua Wang, Dan Western, Jigyasha Timsina, Charlie Repaci, Won-Min Song, Joanne Norton, Pat Kohlfeld, John Budde, Sharlee Climer, Omar H. Butt, Daniel Jacobson, Michael Garvin, Alan R Templeton, Shawn Campagna, Jane O’Halloran, Rachel Presti, Charles W. Goss, Philip A. Mudd, Beau M. Ances, Bin Zhang, Yun Ju Sung, Carlos Cruchaga

## Abstract

Identification of the plasma proteomic changes of Coronavirus disease 2019 (COVID-19) is essential to understanding the pathophysiology of the disease and developing predictive models and novel therapeutics. We performed plasma deep proteomic profiling from 332 COVID-19 patients and 150 controls and pursued replication in an independent cohort (297 cases and 76 controls) to find potential biomarkers and causal proteins for three COVID-19 outcomes (infection, ventilation, and death). We identified and replicated 1,449 proteins associated with any of the three outcomes (841 for infection, 833 for ventilation, and 253 for death) that can be query on a web portal (https://covid.proteomics.wustl.edu/). Using those proteins and machine learning approached we created and validated specific prediction models for ventilation (AUC>0.91), death (AUC>0.95) and either outcome (AUC>0.80). These proteins were also enriched in specific biological processes, including immune and cytokine signaling (FDR ≤ 3.72×10^-14^), Alzheimer’s disease (FDR ≤ 5.46×10^-10^) and coronary artery disease (FDR ≤ 4.64×10^-2^). Mendelian randomization using pQTL as instrumental variants nominated BCAT2 and GOLM1 as a causal proteins for COVID-19. Causal gene network analyses identified 141 highly connected key proteins, of which 35 have known drug targets with FDA-approved compounds. Our findings provide distinctive prognostic biomarkers for two severe COVID-19 outcomes (ventilation and death), reveal their relationship to Alzheimer’s disease and coronary artery disease, and identify potential therapeutic targets for COVID-19 outcomes.

---

Over the past three years, the severe acute respiratory syndrome coronavirus 2 (SARS-CoV-2) that causes coronavirus disease 2019 (COVID-19) has proven to be an extremely contagious and deadly threat to human health. As of June 2022, the virus has caused over 519 million COVID- 19 patients worldwide, leading to 6.2 million deaths and countless other long-term health effects (https://covid19.who.int/). The clinical expression of COVID-19 infection is highly heterogeneous, ranging from asymptomatic or mild symptoms (e.g. mild cold) to critical conditions (e.g. cytokine storm, cardiovascular damage, pneumonia, and death). About 20% of hospitalized COVID-19 patients have become critically ill, requiring oxygen therapy and/or ventilation. More importantly, 20-40% of severe COVID-19 cases (*1*) have an alarmingly low oxygen saturation level (∼50-80%) without respiratory distress, designated as silent hypoxia (*2*). Compared to patients with symptomatic hypoxia, these patients are less likely to seek medical attention resulting in delayed diagnosis and treatment. This distinctive variation of severe COVID-19 manifestation is associated with high death rate. Even in individuals who recover from the short-term symptoms, long-term post-COVID-19 symptoms such as fatigue, cognitive dysfunction, and shortness of breath often persist for many months after recovery from the acute phase of the disease (*3*).

Contrary to earlier studies showing that SARS-CoV-2 was absent from brain (*4*) and cerebrospinal fluid (CSF) (*4*), several recent studies have provided strong lines of evidence that SARS-CoV-2 may directly impact brain function (*5–7*). COVID-19 has negative long-term impacts on the central nervous system (CNS) resulting in a variety of neurological symptoms such as altered taste and smell (observed in ∼15-23% of COVID-19 patients) (*8, 9*), memory decline (“brain fog”; observed in 1 out of 3 patients) (*10*), and Alzheimer’s disease (AD)-like dementia (observed in ∼36% of COVID-19 cases) (*7*). This suggests that SARS-CoV-2 infection can result in brain injury and neurocognitive impairment even in individuals without preexisting neurological conditions. In addition, SARS-CoV-2 affects the myocardium and causes myocarditis and ischemia in 8-12% of COVID-19 admitted patients (*11*). Long-term post- COVID-19 syndrome is shown to be involved with cardiovascular system (elevated cardiac troponin T) (*12*). All these findings together support the long-term impact of COVID-19 infection on CNS dysfunction and cardiovascular system impairment.

Proteomic studies offer a powerful strategy to identify biological pathways implicated on COVID-19 infection and severity (*13*). Similar approaches have been used for aging, cancer, kidney diseases and other viral infections, including Chikungunya virus (*14*) and avian influenza virus (*15*). Several small-scale proteomic studies found that patients infected with SARS-CoV-2 had elevated proinflammatory cytokines including interleukin 6 (IL-6) (*16, 17*), interleukin 1 β (IL-1β) (*16, 18-20*) and tumor necrosis factor (TNF) (*18, 20*). Proteins associated with neutrophil degranulation have also been shown to be upregulated in COVID-19 patients (*21*), suggesting that targeting these pathways in conjunction with antiviral medicine may provide an effective treatment for severe cases (*21, 22*). While these and other proteomic studies have identified proteins significantly associated with COVID-19 status, they have been limited in sample size (<100 participants) or breadth of proteins measured (<200 proteins), reducing power for detecting potentially significant contributing factors to disease outcome and severity.

To understand the pathogenic events linked to COVID-19 outcomes (infection: all cases vs healthy controls; ventilation: cases requiring ventilation vs cases without ventilation; and death: died cases vs survived cases) and identify potential biomarkers and drug targets, we obtained high throughput plasma proteomics (more than 4,300 plasma protein) obtained at the time of hospital admission from two independent cohorts, in order to identify proteomic profiles associated with infection, ventilation or death. The proteins that replicated across studies were then used to create a prediction model for infection (all cases vs healthy controls), ventilation (cases requiring ventilation vs cases without ventilation support), and death (died cases vs survived cases) as well as to identify what biological pathways are dysregulated due to the infection.

## Results

### Study Design

To identify the plasma proteins associated with the COVID-19 outcomes, create prediction models and identify pathways dysregulated with infection, we generated high-throughput proteomic data from 332 COVID-19 patients and 150 controls at Barnes-Jewish Hospital (BJH) and Washington University in St Louis (WUSTL; Figure 1). Among the COVID-19 cases, there were ∼90% symptomatic patients, 93.7% were hospitalized, 46.7% with ICU admission, 24.7% on ventilation, and 19.0% died due to COVID-19 (82 ventilated and 63 died; 44 of the deceased had been ventilated prior to death). COVID-19 patients were 59 years old on average, 58.7% men and 67.8% of African American ancestry (Table 1 and Table S1A). The control group was age-, sex-, and race-matched. Plasma was obtained when admitted to BJH and the event of ventilation and death occurred in 6.8 and 36.2 days respectively after admision on average (Fig. S1). Proteomic data was generated using the SomaScan v4.1 7K panel. After rigorous quality control (QC), a total of 7,055 proteins remained. However, only the 4,301 overlapping proteins with the replication cohort were analyzed in in this study, as there is not possible at this stage to replicate the non-overlapping proteins (See Materials and Methods, and Fig. S2).

**Figure 1:**
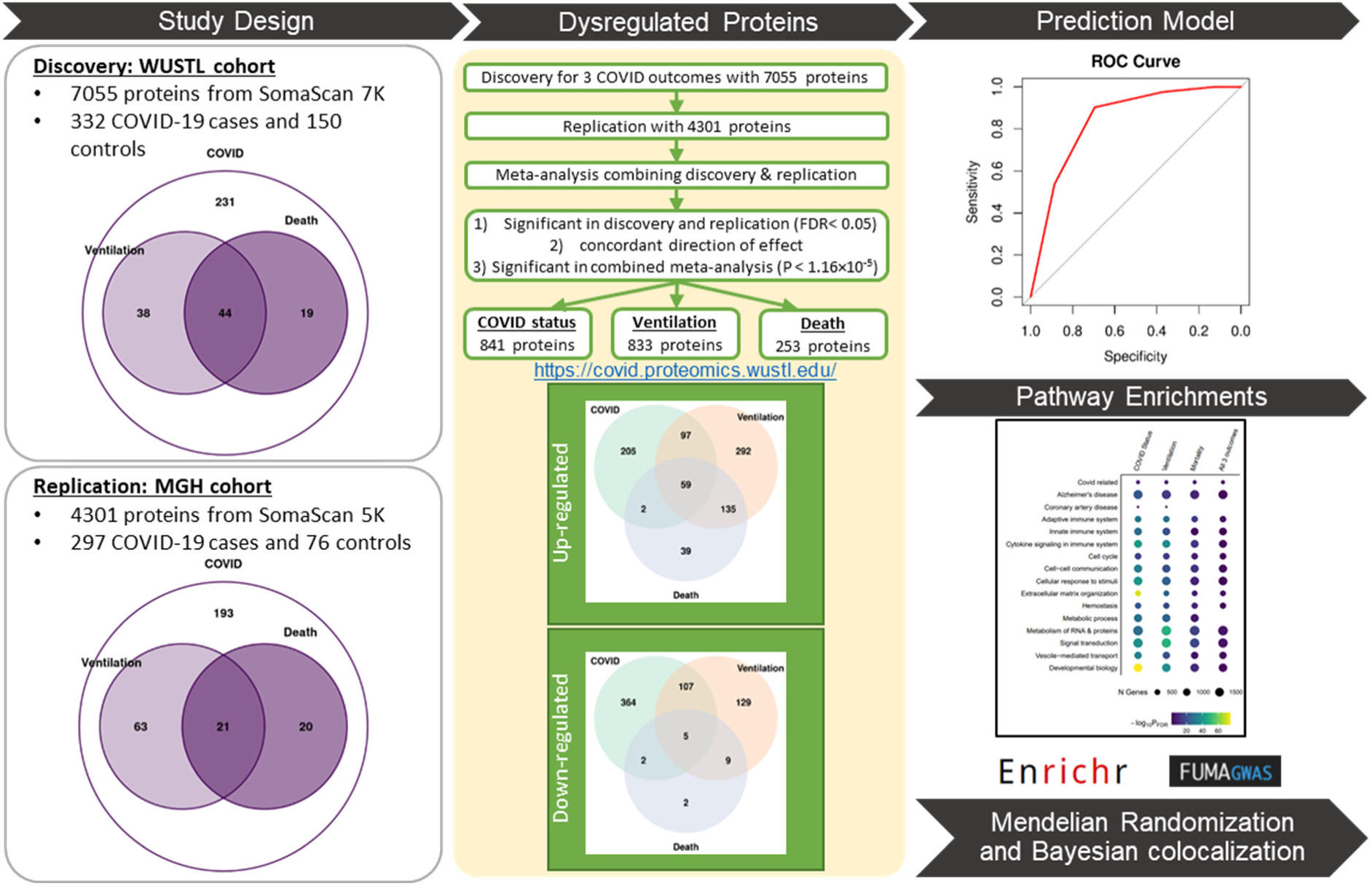
Study overview. In the discovery stage, plasma protein level was measured using SomaScan v4.1 7K (7,055 proteins passed QC) on 332 COVID-19 cases, including 82 patients on ventilation and 63 patients died from SARS-CoV-2 infection, and 150 healthy controls recruited at Washington University in St Louis (WUSTL). Differential abundance analyses for infection, ventilation and death were performed. The publicly available Massachusetts General Hospital (MGH) COVID-19 cohort that includes 4,301 proteins for 297 cases and 76 controls were then used for replication. The replicated proteins were defined by the Benjamini-Hochberg false discovery rate (FDR) < 0.05 in discovery and replication stage, P < 1.16×10^-5^ in meta-analysis and the concordant direction of effect sizes. There were 841 proteins associated with infection, 833 proteins with ventilation and 253 with death, which were used to build and validate prediction models and perform pathway enrichment analyses. Mendelian randomization (MR) was performed using publicly available plasma protein quantitative trait loci (pQTL) of these differential abundant proteins and COVID-19 host genetics initiative (HGI) GWAS summary statistics to identify the causal proteins. For the MR nominated proteins, the genetic region harboring the shared causal variants that driving the causal relationship among them were further tested for Bayesian co-localization analyses. The proteomic data for the discovery cohort be interactively explored in our web portal (www.omics.wustl.edu/proteomics

**Table 1.**
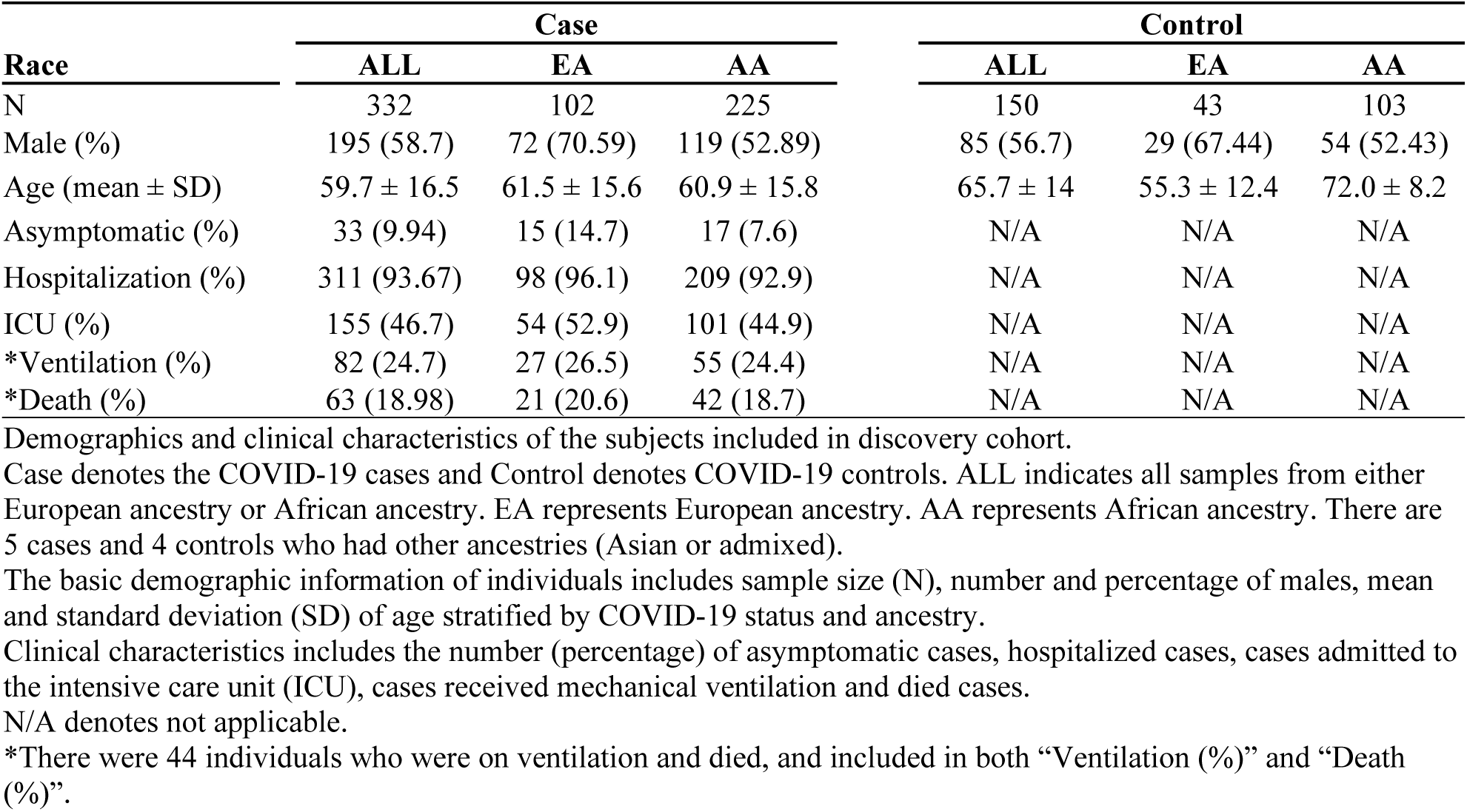
Demographics and clinical characteristics of the discovery WUSTL cohort.

For replication, we leveraged the publicly available proteomics data from 297 COVID-19 cases (84 ventilated and 41 died; 21 of the deceased had been ventilated prior death) and 76 controls recruited at Massachusetts General Hospital (MGH, Table S1B) (*23*). Instead of years, only age range was available for the MGH samples. Most of the controls (44%) were 64-79 years of age. The COVID-19 cases were younger, with 36% in the 49-64 age range. The need for ventilation and death also occurred earlier in this dataset, in 0.5 and 18.2 days, respectively after hospitalization on average (Fig. S1). Proteomic data for the MGH dataset was also generated using the SomaScan platform, but in the 5K panel, where 4,301 proteins passed QC.

We performed differential abundance analysis to identify proteins associated with infection, ventilation and death (the three available COVID-19 outcomes in both datasets). We performed a three-stage study design: 1) Discovery: identification proteins associated with each of these three COVID-19 outcomes at Benjamini-Hochberg false discovery rate (FDR) < 0.05 in the WUSTL cohort; 2) Replication: within the proteins identified on discovery, we identified those proteins that were also associated with COVID-19 outcomes on MGH dataset at Benjamini-Hochberg FDR < 0.05 and in the same direction as in discovery; and 3) Meta-analyses: from the significant proteins remained in replication, we identified those proteins that passed the stringent study-wise Bonferroni threshold (P < 0.05/4,301=1.16×10^-5^, Figure 1) in a fixed-effect meta-analysis, combining discovery and replication results.

Using these significant proteins assayed at patient’s admission to the hospital, we created prediction models not only for infection, but more importantly for ventilation and death, which occurred after blood draw. Prediction models were created using the proteins for all 3 outcomes, as well as ventilation and death-specific models based on the proteins unique for these phenotypes. Prediction models were trained on the WUSTL dataset and validated on the MGH dataset. As the number of proteins in each model were relatively high, we used least absolute shrinkage and selection operator (Lasso) and five-fold cross validation to identify the minimum number of informative proteins for each model.

In addition, pathway enrichment analyses using the proteins that passed multiple test correction on the meta-analyses were performed to identify the biological pathways that were disrupted due to COVID-19 outcomes. In order to identify proteins that are part of the pathological events of COVID-19 outcomes, we performed Mendelian randomization (MR) (see Materials and Methods). We also used causal co-expression network for those proteins associated with COVID-19 approaches to identify proteins that are likely be part of the pathological processes implicated on the infection, and determined if those are also druggable.

### Proteomic signatures of COVID-19 outcomes (infection, ventilation and death)

To identify plasma proteomic signatures for COVID-19 infection (compared to non-COVID-19 infected individuals), we performed association analyses including age, sex, plate and race as covariates (no significant differences were found using surrogate variables instead of plate as shown in Fig. S3). We found 3,236 proteins (1,558 upregulated and 1,678 downregulated) significant after FDR correction in the discovery cohort (Table S2 and Table S5). In the replication cohort, 906 proteins were also associated after FDR correction and in the same direction (Table S5). In the meta-analysis, 841 proteins (363 upregulated and 478 downregulated; 26% out of 3,236; Tables S2 and S5 and Figure 1) were significant after study- wide Bonferroni correction (for the 4,301 proteins; Figure 1 and Tables S2 and S5).

Among the 332 COVID-19 cases from the discovery cohort, 82 individuals needed ventilation in 6.8 ± 7.7 days after hospital admission, and 84 in the replication (time to event: 0.5 ± 1.4; Fig. S1). Using the same three-stages study design as for infection; we identified 2,341 proteins (1,320 upregulated, 1,021 downregulated) associated in the discovery cohort (Table S3 and Table S5), of which 993 were replicated in the same direction, by comparing proteins levels in COVID-19 positive individuals that need ventilation from those that did not need ventilation. In the meta-analysis, 833 proteins (583 upregulated and 250 downregulated; 36% out of 2,341) were significant after Bonferroni correction (Figure 1, Table S3 and Table S5).

There were 63 individuals that died (time to event after hospitalization 36.2 ± 41.2 days) in the discovery cohort and 41 in replication (time to event: 18.2 ± 11.9). We identified 2,101 proteins (1,215 upregulated and 886 downregulated) associated with death in the discovery (Table S4 and Table S5), of which, 297 replicated. A total of 253 proteins (235 upregulated and 18 downregulated; 12% out of 2,101) were significant at Bonferroni corrected threshold in the combined meta-analysis (Figure 1, Table S4 and Table S5).

Finally, we found 64 proteins that were consistently associated with all three COVID-19 outcomes (infection, ventilation and death; Figure 1, Table S5 and Table S6). There were 59 proteins consistently upregulated including Macrophage colony-stimulating factor 1 (CSF-1), Interleukin-1 receptor antagonist protein (IL-1Ra), Leukocyte-specific transcript 1 protein (LST1), Protein kinase C zeta type (PKC-Z), Transforming growth factor beta-1 (TGFB1), Bone morphogenetic protein 10 (BMP10), Angiopoietin-related protein 4 (ANGL4), and Vimentin: i.e. these were upregulated in COVID-19 patients (compared to healthy controls), upregulated in ventilated patients (compared to those who were not) and also upregulated in individuals who died (compared to those who survived; Figure 1). The remaining 5 proteins (Myeloid zinc finger 1 [MZF1], Apolipoprotein A-I [Apo A-I], C-C motif chemokine 22 [MDC], Myosin regulatory light chain 2, atrial isoform [MLRA] and N-acetylglucosamine-1-phosphodiester alpha-N- acetylglucosaminidase [NAGPA]) were consistently downregulated in COVID-19 infection status as well as disease severity.

### Prediction models

Next, we leveraged the proteins that passed the Bonferroni correction in the meta-analyses to create prediction models for the three outcomes. Single protein did not show high and consistent prediction power (Fig. S4), so multi-protein models were created.

#### General prediction models

First, we used the 64 proteins that were associated with all three outcomes to create prediction models for infection, ventilation and death. The area under the curve (AUC) for infection, ventilation and death for the model that included age were higher 0.97 for the three outcomes and in the discovery and replication cohort (Fig. S5A. The positive predictive value (PPV) and negative predictive value (NPV) were estimated based on Youden’s J statistic optimal cut-off using pROC coords function with an optimal cut-off of ∼0.5. The PPV were higher than 94% for all models. The models without age also showed higher AUC, PPV and NPV (Fig. S5B and Table S9), and all models were significantly better than age alone (P<1.1×10^-2^, Table S8 and Table S9).

As the proteins included in the model are too many for a clinically meaningful prediction model and inter-correlated (heatmap shown in Fig. S6 and Table S10), which may lead to overfitting, We obtained a minimum number of proteins with maximum prediction power by using Lasso regression and five-fold cross validation in the discovery cohort. Through this analysis, we identified a subset of 22 proteins (including CSF-1, ANGL4, IL-6) and 10 proteins (including Neurexophilin-1 [NXPH1], Interleukin-1 receptor-like 1 [IL-1 R4], and LST1) that have similar (no significant difference) AUCs, PPV and NPVs (Table S8, Table S9, Fig. S5B) for infection, ventilation and death, when compared to the model with all 64 proteins.

In summary, we identified 64, and a subset of 22 and 10 proteins that can be used to predict individuals infected with COVID-19 and among those the individuals that will need ventilation and death with high accuracy (Table S6), but they cannot be specifically used for prediction of either ventilation only or death only. This limited their application as prognostic tools for clinically monitoring of the progress of COVID-19 patients. A prediction model specific for ventilation (and specific for death) that distinguishes these two outcomes would be informative.

#### Ventilation-specific model

To build a ventilation prediction model, we identified proteins that were: 1) significant in discovery (FDR<0.05) for ventilation, 2) significant in replication (FDR<0.05) for ventilation, 3) significant in the meta-analysis at Bonferronni corrected threshold (P < 1.16 x 10^-5^) for ventilation, 4) concordant direction of effects between discovery and replication, and 5) not significant for death in the meta-analysis (P>0.05). There were 50 proteins (Table S11) that fit these criteria.

As the number of individuals in each group is highly unbalanced, NPV and PPV can be biased. For this reason, NPV and PPV was computed using subsampling with a balanced numbers of individuals in each group and 100 bootstrapped samples (see Materials and Methods; Table S7). The AUCs for the model that included these 50 proteins were 1 for discovery and 0.93 for replication, with PPV>0.88 and NPV>0.93 (Figure 2B, Figure 2C). This model was also significantly better than age alone (P=1.47×10^-9^). In addition, this model was not able to predict death, showing an AUC of 0.53 in discovery and 0.63 for replication, which is even lower than the AUC for age alone (0.55 discovery and 0.88 replication: Figure 2B).

**Figure 2:**
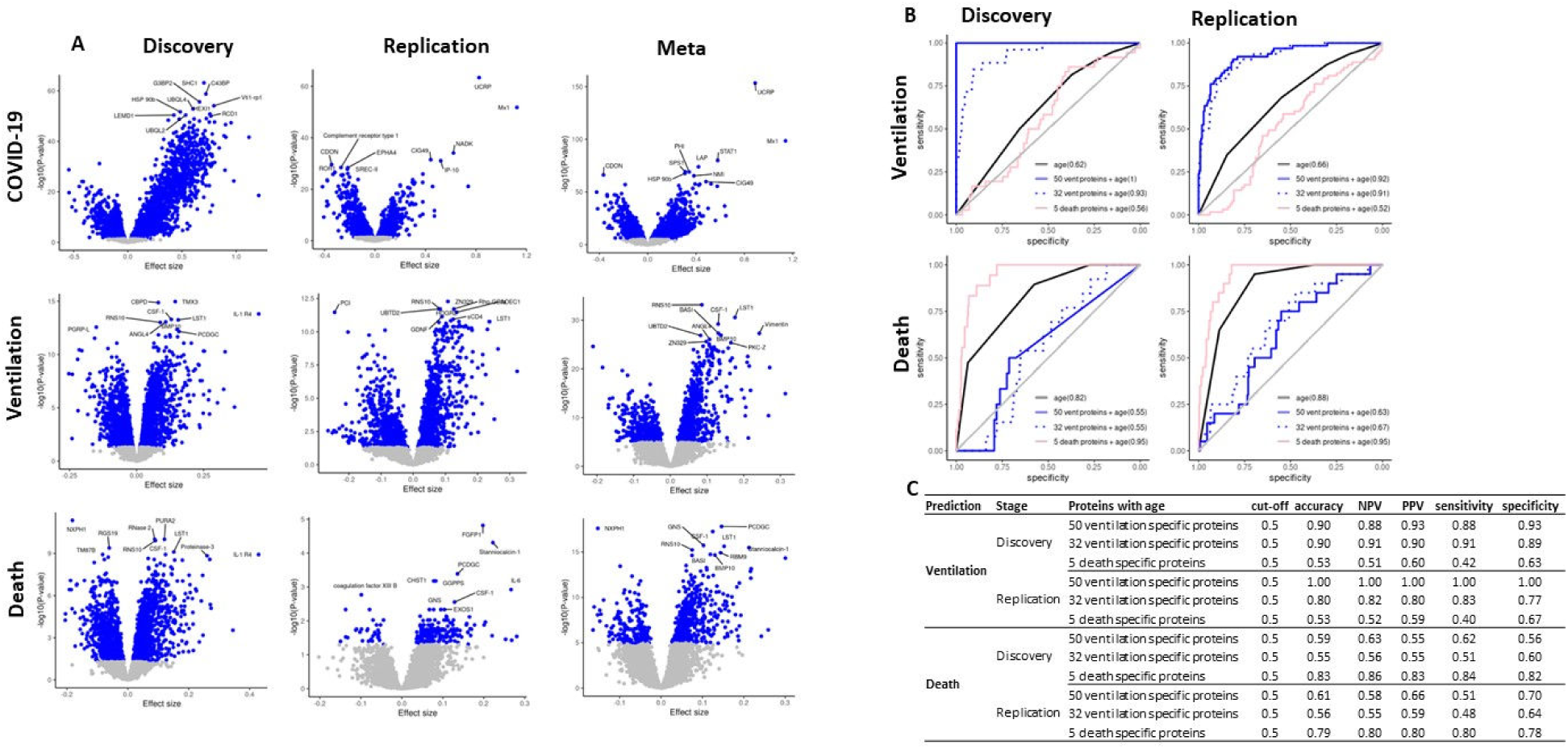
Differentially abundant proteins for COVID-19 infection, ventilation and death and their prediction models. **A**) Volcano plots of differential abundance analyses for COVID-19 infection, ventilation and death in the discovery, replication cohort and in the meta-analysis. Effect size is displayed in the x-axis and –log10(P-value) displayed in y-axis. The blue dots were proteins with the Benjamini-Hochberg FDR<0.05 in discovery and replication, P < 1.16×10^-5^ in meta-analysis. The protein names targeted by the top 10 aptamers are labeled for each of the 6 volcano plots. **B)** The ROC analyses with corresponding AUC were obtained from logistic regression model, which considered each outcome (ventilation and death) as response. The black solid curve included age alone as predictor. The blue solid curve included 50 ventilation specific proteins and age as predictors. The blue dotted curve included Lasso- selected 32 ventilation specific proteins and age as predictors. The pink solid curve included 5 death specific proteins and age as predictors. Y-axis represents sensitivity and x-axis represents specificity. **C)** Table with the performance measures for the 3 prediction models. It includes accuracy, negative predictive value (NPV), positive prediction value (PPV), sensitivity and specificity at 0.5 of Youden’s J statistic. This evaluation used the case-control balanced subsample by selecting age and gender matched or age matched controls in discovery and replication, respectively. For ventilation, the 2 models with 50 and 32 ventilation specific proteins outperformed, providing ∼77-100% prediction values. For death, the model with 5 death specific proteins performed better, showing ∼78-86% prediction values.

Similar to the general prediction model, we used LASSO to identify a subset of 32 proteins that provided a similar performance as the model with the 50 proteins (Table S11). The model with the 32 proteins (Figure 2B and Figure 2C) had AUC higher than 0.91 and PPV and NPV higher than 0.80 for ventilation, while showing low prediction power for death (AUC< 0.67). Results were similar without age in the model for the above protein sets (Fig. S7). These results indicate that these 50 (and the subset of 32) at hospital admission can predict need for ventilation significantly better than age alone, with high sensitivity and specificity.

#### Death-specific model

To create death-specific models we used a similar approach to that of ventilation. We identified proteins that were: significant in discovery (FDR<0.05) and replication (FDR<0.05), the meta- analysis (Bonferroni corrected P < 1.16×10^-5^), concordant direction of effects for death, and 5) not significant for ventilation in the meta-analysis (P>0.05). This led to the identification of 5 proteins: Ankyrin repeat and SOCS box protein 9 (ASB9), Endothelial cell-specific molecule 1 (Endocan), Transcription factor jun-D (jun-D), Pikachurin (EGFLA), Glutathione S-transferase Pi (Table S11).

The AUCs for death using these five proteins was 0.95 in both discovery and replication, with high PPV (0.83 in discovery and 0.80 in replication) and high NPV (0.86 in discovery and 0.80 in replication). This model was significantly higher than AUC for age alone (P=2.29×10^-3^ and P=1.28×10^-2^). At the same time, the AUCs of these five proteins for ventilation were very low (<0.56), with ∼0.6 PPV and ∼0.5 NPV. Again, compared with the poor performance of predicting death using ventilation specific proteins, these death specific proteins showed promising results with accurate prediction, indicating that these proteins can identify individuals that would be at higher risk of death after COVID-19 infection.

### Proteins associated with COVID-19 infection, ventilation and death are enriched in immune- related, Alzheimer’s disease and cardiovascular disease pathways

To identify the functional pathways and biological processes involved in SARS-CoV-2 outcomes, we performed enrichment analysis for the significant proteins in each outcome (841 for COVID-19 status, 833 for ventilation and 253 for death) and the 64 proteins in common for all 3 outcomes using FUMA GWAS (*24*) and Enrichr (*25*), which includes a specific “COVID- 19 Related Gene Sets 2021” pathway.

We found an enrichment for the “genes down-regulated by SARS-CoV-2 infection”(*26*), a COVID-specific pathway that was created based on cell models after COVID-19 infection (GEO accession GSE147507). This pathway was enriched for all 3 outcomes: infection, ventilation, and death (FDR = 2.9×10^-5^, 3.9×10^-2^, and 4.6×10^-2^, respectively). There were many other pathways enriched for immunologic signatures including adaptive (FDR<1.2×10^-15^), innate (FDR<5.0×10^-4^), B cells response from influenza vaccine (FDR < 7 ×10^-6^) or cytokine signaling in immune system (FDR<3.7×10^-14^; Figure 3A, Table S13). However, we also identified an enrichment for pathways that are not related to the immune response to a virus infection. They included Alzheimer’s disease (FDR<5.5×10^-10^) and coronary artery disease (FDR<4.6×10^-2^), suggesting that COVID-19 infection may impact pathways and organs beyond what would be expected.

**Figure 3.**
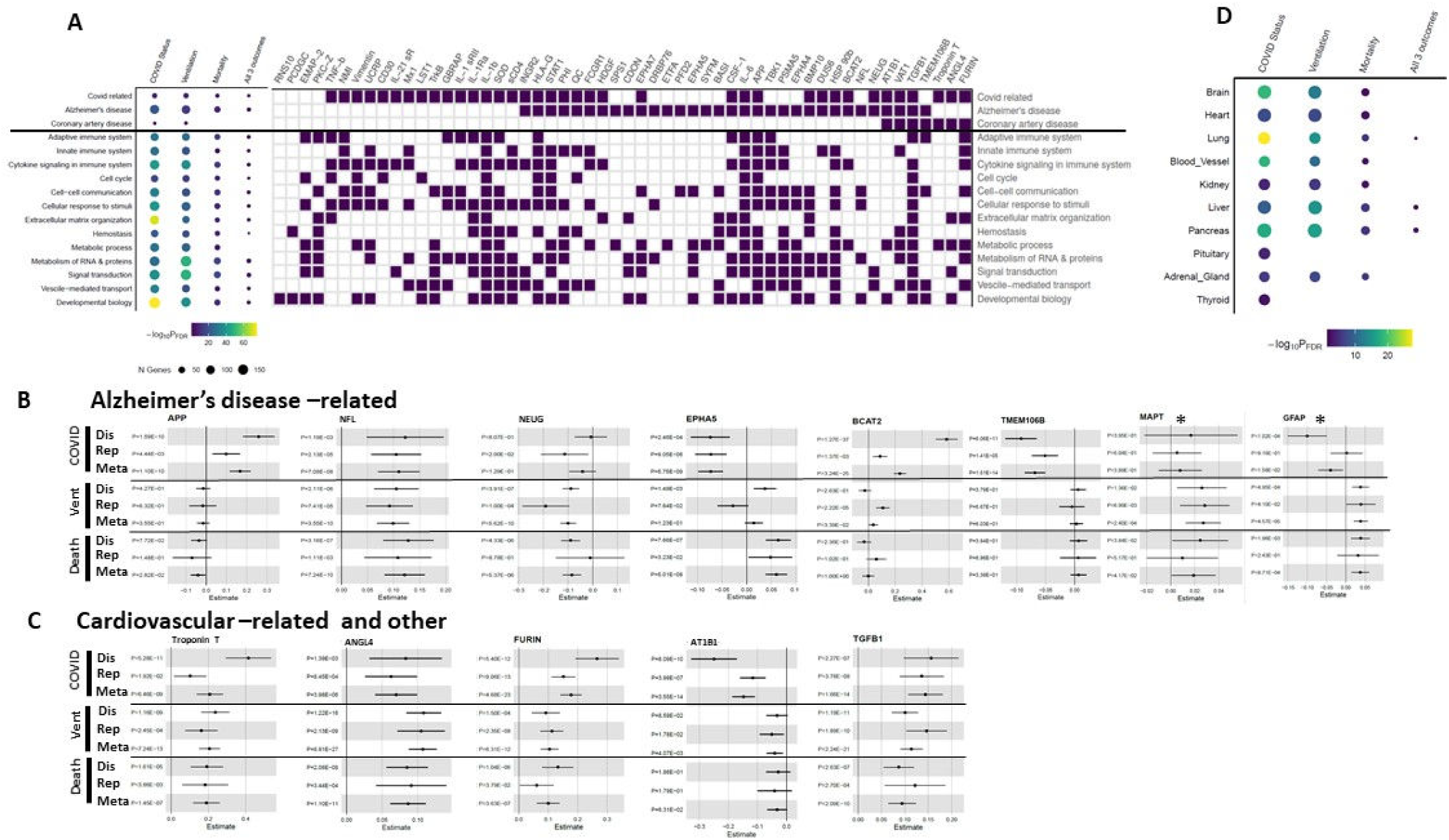
Pathway enrichment analyses using the robustly identified proteins. **A)** Pathway enrichment analyses of proteins identified on the meta-analyses identified 16 different pathways. Size in the dot chart corresponds to the number of identified proteins and color corresponds to the FDR corrected significance. Several differentially abundant proteins (IL-6, IL-1b, IL-1Ra, and STAT1, among others) for the thre outcomes were enriched in COVID-19 related pathways. Proteins were significantly enriched for Alzheimer’s disease (AD) pathway (FDR=6.84×10^-22^ for COVID-19 infection; FDR=1.19×10^-14^ for ventilation; FDR=5.46×10^-10^ for death). Enrichment of COVID-19 infection (FDR=0.046) and ventilation (FDR=0.019) was observed for coronary artery disease. **B)** Forest plots of AD-related proteins: amyloid precursor protein (APP), Neurofilament light polypeptide (NFL), Neurogranin (NEUG), Ephrin type-A receptor 5 (EPHA5), Branched-chain-amino-acid aminotransferase, mitochondrial (BCAT2), Transmembrane protein 106B (TMEM106B), Microtubule-associated protein tau (MAPT*), Glial fibrillary acidic protein (GFAP*\**). *: showed a nominal significance (P<0.05). **C)** Forest plots of cardiovascular-related proteins: Troponin T (cardiac muscle), Angiopoietin-related protein 4 (ANGL4), FURIN, Sodium/potassium-transporting ATPase subunit beta-1 (AT1B1) and Transforming growth factor b1 (TGFB1). **D)** Tissue specificity enrichment analyses via FUMA for 30 tissues in GTEx v8 using the robustly identified proteins. In addition to enrichment for lung, we found enrichment for brain (P=2.03×10^-19^, 4.79×10^-13^, and 6.3×10^-3^ for COVID-19 infection, ventilation and death, respectively) and enrichment for heart (P=2.81×10^-8^, 6.02×10^-8^, and 3.03×10^-3^ for COVID-19 infection, ventilation and death, respectively).

#### Cytokine and immune related pathways

Among key immunopathological consequences of severe COVID-19, a cytokine storm, characterized by elevated levels of pro-inflammatory mediators, is shown to be a central feature, as reviewed (*22*). The proteins identified and replicated across all three outcomes were significantly enriched in cytokine signaling in immune system (GO:0034097; FDR = 1.99×10^-42^, 2.3×10^-38^ and 3.72×10^-14^ for infection, ventilation and death, respectively) including Interleukin- 1 receptor antagonist protein (IL-1Ra), IL-6, Interleukin-21 receptor (IL-21 sR), TGFB1, Vimentin, Hepatoma-derived growth factor (HDGF), and CSF-1. The innate (R-HSA-168249; FDR ≤ 5.03×10^-4^) and adaptive immune system (GO:0006954; FDR ≤ 1.23×10^-15^) were also highly significant in these analyses. Several proteins (Phospholipase A2, membrane associated [PLA2G2A], C-reactive protein [CRP], Amyloid beta A4 protein, ADP-ribosyl cyclase/cyclic ADP-ribose hydrolase 2 [BST1]) were shared across these three immune-related pathways.

IL-1Ra inhibits the activities of IL-1A and IL-1B, and modulates IL-1 related immune and inflammatory responses of acute infection. Anakinra, a FDA approved immunosuppressive drug for rheumatoid arthritis, deficiency of IL-1A, demonstrated improved overall survival and invasive ventilation-free survival for acute respiratory distress syndrome patients associated with COVID-19 (*27*). Consistent with high IL-6 levels in all 3 COVID-19 outcomes observed in this study (P < 2.7×10^-11^), IL-6 levels are shown to be increased in COVID-19 severe cases compared with those in non-severe condition (*28*). IL-21 and IL-21sR (P < 1.1×10^-9^) are part of both innate and adaptive immune responses, playing an important role in viral infections, and the combination therapy of IL-15 and IL-21 for COVID-19 was warranted (*29*). TGFB1 has been shown to suppress the type I interferon (IFN) response in alveolar macrophages (*30, 31*). We also observed lower levels of type I interferon receptors (IFN-a/b R1; P=8.13×10^-17^), IFNA5 (P=1.06×10^-9^), IFNA2 (P=5.10×10^-9^), IFNB1 (P=1.99×10^-6^), and IFNA14 (P=4.42×10^-6^) in COVID-19 cases. The TGF-b dominated immune response from COVID-19 was observed in single cell transcriptome levels from plasmablasts (*32*). We found higher TGFB1 levels in COVID-19 patients (P < 4.0×10^-11^) across all three outcomes. Macrophage colony-stimulating factor 1 (CSF-1) is a cytokine and involved in innate immunity and inflammatory process, regulating survival, proliferation and differentiation of macrophages and monocytes. Higher CSF-1 levels have been reported in serum of COVID-19 patients and proposed to be a potential target for COVID-19 (*33*). We found significantly higher CSF-1 levels in COVID-19 patients (P < 4.1×10^-17^) across all three outcomes. Vimentin, a type III intermediate filament protein, along with microtubules and actin microfilaments, makes up the cytoskeleton which is responsible for maintaining the integrity of the cytoplasm. Vimentin has been reported as a co-receptor for SARS-CoV-2 (*34*), involved in viral replication in cells, and proposed as a target for COVID-19. We found that this protein was significantly higher in COVID-19 patients (P < 1.5×10^-14^) across all three outcomes. HDGF has been shown to be significantly upregulated in transcriptome of liver autopsy samples of severe COVID-19 patients (*35*). We found high HDGF levels in our COVID-19 patients across all three outcomes (P < 4.1×10^-14^).

#### Alzheimer’s disease (AD) related pathways

Central nervous symptom (CNS) damage can be present early in the course of COVID-19. Reiken et al (*36*) reported Alzheimer’s-like signaling including tau hyperphosphorylation and leaky ryanodine receptor/calcium release channels on the endoplasmic reticuli in brains of COVID-19 patients. Small-scale studies found that some neurodegeneration-related biomarkers such as NFL were elevated in COVID-19 patients (*37*), suggesting a potential impact of COVID- 19 on brain health.

Several proteins associated with COVID-19 outcomes in our meta-analyses are part of the Alzheimer’s disease (AD) pathway (*38*) (Molecular Signatures Database M12921; FDR = 6.84×10^-22^, 1.19×10^-14^, and 5.46×10^-10^ for infection, ventilation and death, respectively). Proteins that are part of this pathway include Neurofilament light polypeptide (NFL), Neurogranin (NEUG), Ephrin type-A receptor 5 (EPHA5), Branched-chain-amino-acid aminotransferase, mitochondrial (BCAT2), Transmembrane protein 106B (TMEM106B), Microtubule-associated protein tau (MAPT), Glial fibrillary acidic protein (GFAP; Figure 3A). We found association of MAPT with ventilation and association of GFAP with ventilation and death at a nominal significance.

Amyloid precursor protein (APP) is a well known AD gene and was significantly higher in COVID-19 cases (P=1.1×10^-10^). Genetic variants in transmembrane protein 106B (*TMEM106B*) are associated with risk for Parkinson disease (PD) (*39*), Frontotemporal dementia (FTD) (*40*) and AD (*41*). Our group has also demonstrated that *TMEM106B* is associated with neuronal survival in AD (*42*). Recent studies also suggest that TMEM106B is a proviral host factor for SARS-CoV-2 (*43*). TMEM106B is a component of the lysosome. The SARS-CoV-2 virus hijacks the lysosome to exit the cells via ORF3a and NSP6 (*44*). Lower plasma TMEM106B protein levels reflecting disruption of normal lysosome function were found in COVID-19 patients (P=1.51×10^-14^). EPHA5 has been reported to be associated with early-onset familial AD (*45*), and our group also leveraged CSF pQTL, colocalization and MR to demonstrate that EPHA5 is a causal protein for AD (*46*). We found that EPHA5 was significantly lower in COVID-19 patients (P=5.75×10^-9^). Mutations and common variants in *MAPT* have been associated with FTD (*47*), PSP (*48*) and AD (*49*). We found that higher levels of MAPT were associated with higher risk of ventilation (P=2.4×10^-4^) at a nominal significance.

Several well known biomarkers for AD and neurodegeneration (including neurogranin, NFL and GFAP) were also altered due to infection or associated with ventilation and death. Neurogranin, a cognitive biomarker in CSF and blood exosomes for AD (*50*), was significantly lower in plasma of ventilated COVID-19 patients (P=5.62×10^-10^). NFL reflects neuronal death and is one of the promising blood biomarkers for AD and neurodegeneration in general (*51*). NFL was significantly increased (P<7.08×10^-8^) in COVID-19 across all 3 outcomes. We found that GFAP, a biomarker for astroglial pathology in neurological diseases (*52*), was higher in COVID-19 patients that needed ventilation or died (P<1.58×10^-2^) at a nominal significance.

In summary, we found that multiple proteins that are part of the causal pathway of AD or are biomarkers for AD and neurodegeneration were altered in COVID-19 patients, particularly those with poor outcomes. We also performed tissue specificity enrichment test and found that the proteins associated with COVID-19 infection and outcomes were enriched for brain-specific proteins (P=2.03×10^-19^, 4.79×10^-13^, and 6.3×10^-3^ for COVID-19 infection, ventilation and death respectively; Figure 3D, Table S14).

#### Coronary artery disease (CAD) related pathways

It has been shown that SARS-CoV-2 infection can increase the risk of cardiovascular disease. Even a mild case of COVID-19 is reported to increase risk of cardiovascular disease, independent of age and smoking history for at least a year after diagnosis (*53*). Our analyses indicate that the proteins associated with COVID-19 outcomes are enriched for coronary artery disease (CAD) related pathways (FDR=0.046 for COVID-19 infection, and FDR=0.019 for ventilation; Figure 3A, Table S13). Proteins that are part of this pathway include troponin T, angiopoietin-related protein 4 (ANGL4), FURIN, sodium/potassium-transporting ATPase subunit beta-1 (AT1B1) and transforming growth factor b1 (TGFB1). Troponin T is a protein integral to cardiac muscle and a known plasma biomarker for myocardial infarction (*54*). We fount significantly higher Troponin T levels in COVID-19 patients (P=6.46×10^-9^; Figure 3C) and those requiring ventilation (P=7.24×10^-13^; Figure 3C). Genetic studies have confirmed that loss-of-function variants of *ANGL4* significantly reduced risk of CAD (*55*), and our analyses found significantly increased ANGL4 levels in COVID-19 infection (P=3.98×10^-6^; Figure 3C), ventilation (P=8.91×10^-27^; Figure 3C) and death (P=1.1×10^-11^; Figure 3C). Genetic variants that increase the expression of *FURIN* has been associated with coronary artery disease (*56*), and we found significantly higher FURIN levels in COVID-19 infection (P=4.68×10^-23^) and ventilation (P=6.31×10^-12^). *ATP1B1* is a causal gene for CAD (*57*), and we found significantly lower ATP1B1 levels in COVID-19 patients (P=3.55×10^-14^; Figure 3C). Our tissue-specific enrichment analysis also supports that COVID-19 affects heart-specific proteins (enrichment P=2.81×10^-8^, 6.02×10^-8^, and 3.03×10^-3^ for COVID-19 infection, ventilation and death, respectively; Table S14 and Figure 3D)

### Identification of causal and potential therapeutic targets

Pathway analyses point to specific biological pathways that are altered due to COVID-19 infection and that are associated with specific outcomes, such as ventilation and death, but those analyses cannot differentiate the causal proteins from those that are due to secondary changes. In order to identify potential causal pathways, we performed MR using plasma pQTL as instrumental variables for those proteins that were associated with infection, ventilation and death, as well as performed Bayesian network analyses to identify key drivers and potential targets for known therapies.

#### Mendelian randomization (MR) analyses and co-localization analyses

A recent GWAS including 49,562 COVID-19 patients identified 8 loci associated with infection, 6 loci associated with critical illness, and 9 loci for hospitalization after COVID-19 infection (*58, 59*). In parallel to this, there was a recent plasma pQTL study based on 4,907 plasma proteins in 35,559 Icelanders (*60*), which included 1,437 proteins that we found associated with COVID-19 infection, ventilation or death (total n=1,449). We integrated the COVID-19 GWAS results in European population and pQTLs of the 1,437 plasma proteins, and performed two-sample MR analyses to identify the causal plasma biomarkers.

We found that BCAT2 and GOLM1 were significant in our MR analyses after multiple test correction (Bonferroni corrected threshold= 0.05/1,437=3.48×10^-5^) for COVID-19 infection. For BCAT2, four independent SNPs were used as instrument variables (IV; Table S17), using the inverse variance weighted (IVW) approach (P=2.13×10^-5^). Because MR results can be affected by pleiotropy, we performed several sensitivity analyses to test for any evidence of pleiotropy (Cochran’s Q heterogeneity, Egger intercept, and MR-PRESSO). We found no significance in any of these tests, indicating that our MR results for BCAT2 were not driven by pleiotropy. One of the four IVs used in this analysis is a *cis*-pQTL for BCAT2 (rs11548193; P=3.09×10^-69^; Figure 4D, Figure 4E) associated with COVID-19 infection (P=1.09×10^-6^). MR analysis including only this region was also significan (P=0.001), supporting that the results were not driven by pleiotropy. The effect estimate for MR was also in the same direction as in our differential abundant analyses, with higher BCTA2 levels associated with infection and worse outcomes. BCAT2 is a branched chain aminotransferase found in mitochondria. We also performed Bayesian colocalization analysis and found strong evidence of colocalization (posterior probability PP.H4 = 0.994; Figure 4C), indicating a presence of functional genetic variants affecting both BCAT2 and COVID-19 infection. Our results along with the evidence from the capture Hi-C omnibus gene score (COGS) (*61*) indicate that BCAT2 is a key plasma biomarker for COVID-19 and its potential application as a drug target needs further evaluation.

**Figure 4.**
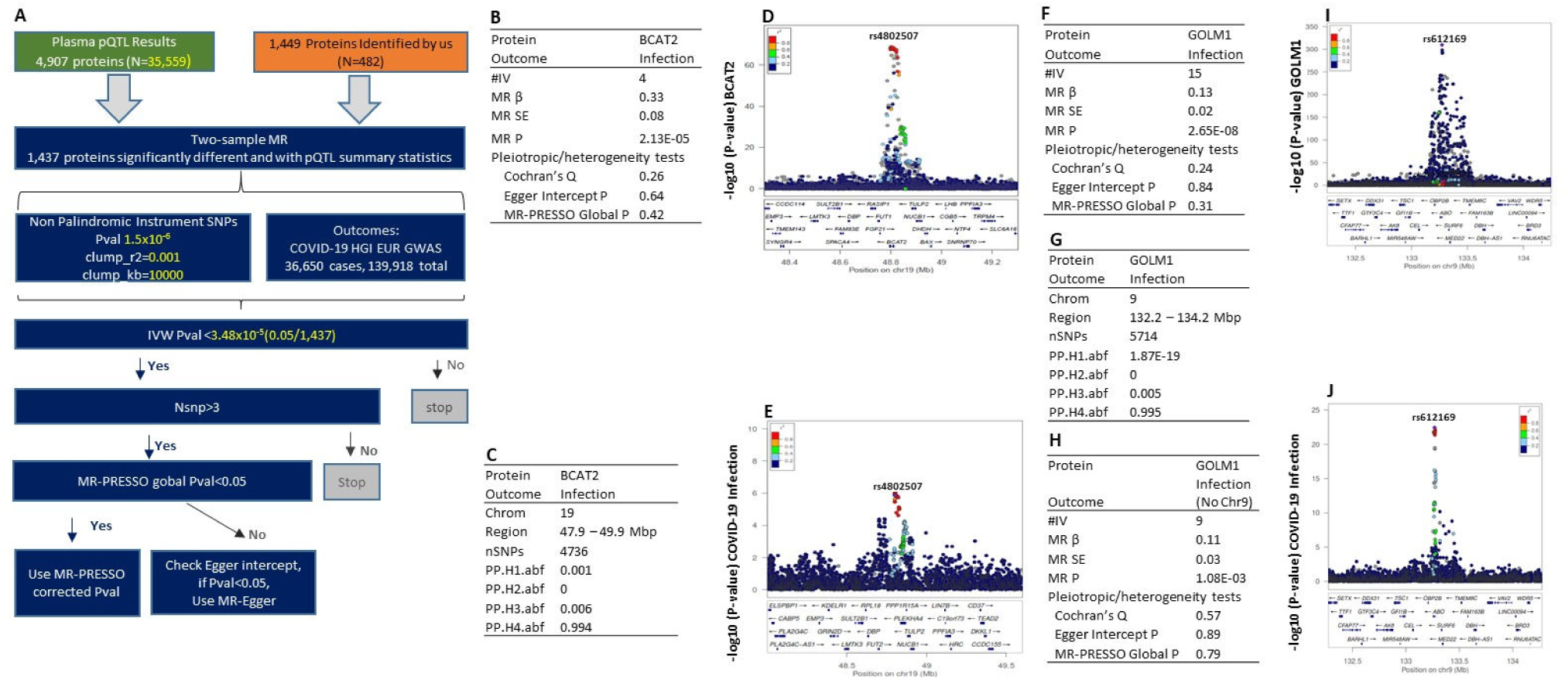
Two-sample Mendelian randomization (MR) workflow and results nominating BCAT2 and GOLM1 as causal candidates for COVID-19 infection. **A)** Two sample MR were performed to identify the causal plasma biomarkers for COVID-19 infection using pQTL summary statistics of each of 1,437 differential abundant proteins identified by our analyses and GWAS of COVID- 19 infection from HGI as outcomes. The independent non-palindromic pQTL with P<1.5×10^-6^ were selected using clumping r2=0.001 and clumping kb=10000 as instruments. The inverse variance weighted method (IVW) was the primary methods of MR analyses. The criteria of significance for MR is P<3.48×10^-5^. If the number of instrument variables were larger than 3, MR pleiotropy residual sum and outlier (MR-PRESSO) was used to detect horizontal pleiotropy. If MR-PRESSO global P<0.05, we corrected horizontal pleiotropy via outlier removal. If MR-PRESSO global P>0.05 and Egger intercept P<0.05, P from MR-Egger method will be used for MR analyses. **B)** MR analyses identified BCAT2 as a causal plasma biomarker for COVID-19 infection. The MR effect estimate (β) is in the same direction as our differential abundant analyses. #IV: number of independent instrument variants: MR β: MR effect estimate; MR SE: MR standard error of estimated effect. **C)** Bayesian colocalization analysis found a presence of the functional variants who have driven this causal effect in chromosome 19. PP.H1.abf: Only BCAT2 has a genetic association in the region. PP.H2.abf: Only COVID-19 infection has a genetic association in the region. PP.H3.abf: Both BCAT2 and COVID-19 infection has a genetic association in the region, but with different causal variants. PP.H4.abf: BCAT2 and COVID-19 infection shared a single causal variant in the region. **D)** Locuszoom plot of rs4802507 in chromosome 19 for pQTL of BCAT2. **E)** Locuszoom plot of rs4802507 in chromosome 19 for GWAS of COVID-19 infection. **F)** MR analyses also identified GOLM1 as another causal plasma biomarker for COVID-19 infection. The MR β is in the same direction as our differential abundant analyses. **G)** Baysian colocalization analysis found a presence of the functional variants in chromosome 9. PP.H1.abf: Only GOLM1 has a genetic association in the region. PP.H2.abf: Only COVID 19 infection has a genetic association in the region. PP.H3.abf: Both GOLM1 and COVID-19 infection has a genetic association in the region, but with different causal variants. PP.H4.abf: GOLM1 and COVID-19 infection shared a single causal variant in the region. **H)** Sensitivity MR analyses by removal of variants at chromosome 9 ABO locus for GOLM1, MR P is still significant. **I)** Locuszoom plot of rs612169 in chromosome 9 for pQTL of GOLM1. **J)** Locuszoom plot of rs612169 in chromosome 9 for GWAS of COVID-19 infection.

We found GOLM1 as another potential causal plasma protein for COVID-19 infection (MR IVW P=2.65×10^-8^; Figure 4F). A total of 15 instrument variables were included in the analyses (Table S19). One of the pQTL used as an IV in the analysis was located at chromosome 9 near rs612169 (intronic variant of ABO). Since the ABO locus is a well-known pleiotropic locus, we further performed a sensitivity analyses by removing ABO locus and found that MR was still significant (P=1.08×10^-3^). We also found no evidence of pleiotropy. The effect estimate was in the same direction as in the differential abundant analyses, with higher protein level associated with COVID-19 infection. Our Bayesian colocalization analysis showed presence of common functional variants (PP.H4 = 0.995) within ABO locus affecting both GOLM2 and COVID-19 infection (Figure 4G). These analyses indicate that GOLM1 is another causal plasma protein for COVID-19 infection.

#### Causal network analyses

Gene co-expression network analysis is commonly used to identify and prioritize potentially functional genes and clusters of genes contributing to disease. However, many of these techniques are limited by prior assumptions on the number of clusters or set thresholds for determining co-expression. We used MEGENA (*62*), a package to perform co-expression analysis that avoids this limitation. MEGENA uses a Bayesian approach to create causal models to dissect genetic pathways and provide mechanisms implicated in disease. Using MEGENA, we built co-expression networks based on the robustly identified proteins associated with COVID- 19 outcomes, and distinguish clusters of correlated proteins. Based on these clusters, MEGENA identified highly-connected hub proteins that are potentially novel key drivers of COVID-19 infection and severity. These hubs were chosen as the focus of our analyses.

We used as input all differentially abundant proteins across three outcomes (COVID-19 infection, ventilation, and death), totaling 1,449 unique proteins. We ran MEGENA using the hub proteins identified in the combined runs to identify clustering of the hubs. For the 1,449 proteins that were significant from any of three outcomes, we identified 141 key (hub) proteins through MEGENA (Table S21). These proteins were used as input to MEGENA to identify clusters, of which eight were identified; 47 proteins were not assigned to a cluster (Figure 5). Of these 141, at least 20 have links to immune function according to GeneCards (*63*). At least 15 are kinase-related genes, and evidence supports the use of kinase inhibitors in COVID-19 treatment (*64*). An additional 21 are involved in the extracellular matrix or cellular adhesion, suggesting potential importance in the virus-cell interface. We matched the identified hub proteins to pathways enriched for differentially abundant proteins (Table S21). The Gene Ontology sets containing the most hub proteins are proteolysis (31 hub proteins), proteinp (27), positive regulation of signaling (27), Blalock Alzheimer disease up (25), and Naba matrisome (25). Dysregulation of protein phosphorylation has been identified in Vero E6, supporting a link between these hub proteins and COVID-19 (*65*).

**Figure 5.**
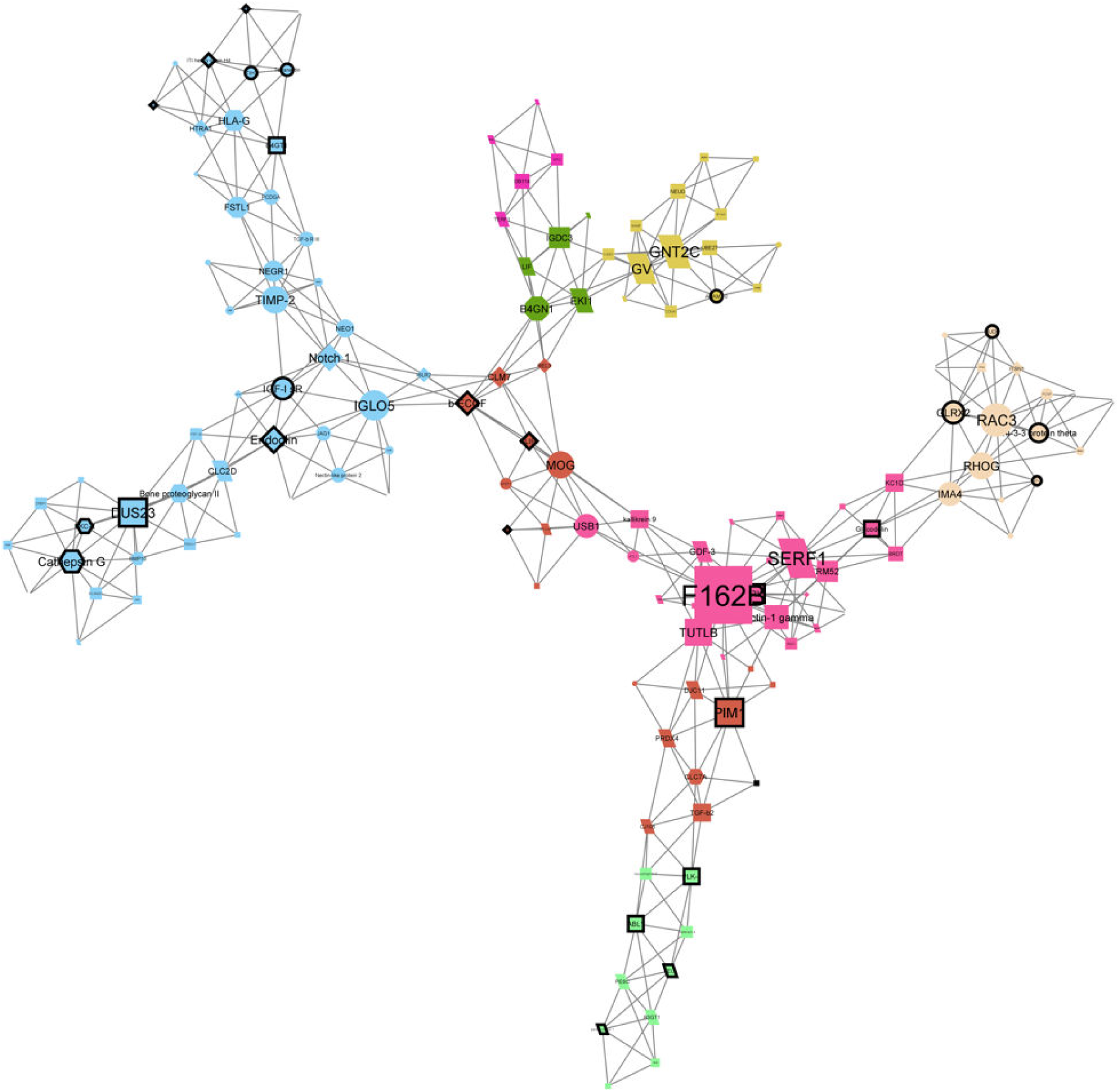
A network plot containing 141 hub proteins identified from combined analysis of 1,449 differentially abundant proteins. MEGENA network plot produced in Cytoscape using 141 hub proteins identified from MEGENA analysis of all differentially abundant proteins across the three COVID-19 oucomes. Colors represent clusters identified by MEGENA. Nodes with dark outlines represent proteins targeted by drugs according to DrugBank. Node shape corresponds to the analyses where that protein was significant. Protein nodes shaped as circles are significantly dysregulated only in COVID-19 infection (case vs control) status, triangles in infection and death (patients who died vs who did not), diamonds in infection and ventilation (patients on ventilation vs without ventilation), hexagons in all three, octagons in death, rectangles in ventilation, and parallelograms in ventilation and death.

Among the 141 hub proteins, we found that 35 have associated drugs in the DrugBank database and 19 of them have a drug with previous links to antiviral activity (Figure 5, Table S21). One drug, Fostamatinib, was found to target 5 hub proteins (ABL1, RIPK2, PIM1, PLK-1, and PAK7). Fostamatinib is a Tyrosine kinase inhibitor usually used to treat chronic immune thrombocytopenia (*66*). Studies support it for treatment of acute respiratory distress syndrome in COVID-19 (*67*) and a stage 2 clinical trial found it safe and showed evidence of better outcomes for COVID-19 patients compared to placebo, suggesting its standard use for care (*68*). In addition, PRIO (encoded by *PRNP*) and MK12 (encoded by *MAPK12*) are both targeted by tetracyclines. These antibiotics have been used to treat numerous types of infection, including pneumonia, cholera, and syphilis (*69*). Tetracyclines are also reported to protect against HIV (*70*). They may be able to act through multiple pathways, including inhibition of RNA replication or downregulation of the NF-κB pathway, in order to decrease severity of COVID-19 (*69*). A network plot of all 1,449 proteins produced using MEGENA and Cytoscape is included as Fig. S10.

## Discussion

Deep plasma proteomic profiling of COVID-19 patients can be instrumental not only to identify the biological processes implicated in response to COVID-19, but also to discover models that can predict the progression to future severe outcomes such as ventilation and death. Here we generated and analyzed deep proteomic data from two large and well characterized independent cohorts to identify proteins dysregulated due to COVID-19 infection and associated with later need of ventilation and death. We used a three-stage study design (discovery, replication and meta-analysis) with stringent multiple test correction to identify a robust set of associated proteins. Our analyses identified 1,449 proteins that replicated and passed Bonferroni correction. There were 841 proteins associated with COVID-19 infection, 833 proteins with ventilation and 253 proteins for death.

Several methods have been used to try to identify individuals with bad outcomes after COVID- 19 infection. They include the Charlson Comorbidity Index Score (CCIS) (*71*), the Sequential Organ Failure Assessment (SOFA) score or the Acute Physiology and Chronic Health Evaluation (APACHEII). However, all these methods showed limited performance in predicting the severe COVID-19 outcome (*72–74*). As a promising strategy, a recent proteomic study identified plasma prognostic biomarkers including several markers of neutrophil activation (resistin, lipocalin-2, hepatocyte growth factor, interleukin-8, and granulocyte colony-stimulating factor), which precede the onset of critical illness with high discriminatory power for detecting critical illness (*75*). However, this study only tested 78 immunologic function related proteins to distinguish control, non-ICU COVID-19 and ICU COVID-19 samples and was lacking the extensive evaluation of classification performance. There was another study identifying a set of biomarkers (AGT, CRP, SERPINA3, and PLG) from 1,169 plasma proteome, predicting the need for invasive mechanical ventilation with 0.98 of AUC (*76*). This study was also limited by relative small sample size (139 COVID-19 patients). Therefore better and more robust prediction models are needed.

Plasma proteins dysregulated in COVID-19 cases or associated with bad outcomes can be leveraged to create prognostic biomarkers to identify individuals at high risk of severe illness who might need intensive management. Using the 64 proteins associated with infection, ventilation and death, we created a prediction model that can differentiate individuals that have an ongoing COVID-19 infection, and within the COVID-19 positive individuals (AUC>0.97) those that will need ventilation (AUC> 0.94) or that will die (AUC=1; Fig. S6). Through machine learning, we identified 10 proteins that showed similar predictive power as those 64 could be implemented in a clinical test. This model predicts all three outcomes (infection, ventilation, and death) with high accuracy, but it does not distinguish between more severe outcomes. We next created prediction models specific for ventilation and death. From the identified proteins, we created a robust prediction model specific for ventilation using the 50 proteins only associated with ventilation (and not with infection or death), or a subset of 32 proteins that may be easier to measure in a hospital setting. This model can accurately distinguish high-risk COVID-19 patients that will require mechanical ventilation 0.5-6.8 days before the event. This predictive model showed highly predictive power (AUC >0.9; Figure 2) in both discovery and replication for identifying individuals that need ventilation, and did not predict individuals that will die from COVID-19. Similarly, a robust prediction model using 5 unique death-associated proteins demonstrated significantly high AUC (0.95; Figure 2) to identify patients that will die after 18-36 days from blood collection. This model is specific to death and does predic ventilation or infection. These three sets of proteins can be used as prognostic biomarkers to guide the allocation of medical resources to COVID-19 cases at high risk for severe illness.

The proteins associated with COVID-19 infection and outcome can also be essential to identify the dysfunctional biological processes in pathways and nominating potential causal proteins implicated in COVID-19. One of the most significant pathways (*26*) is a newly defined set of genes that were identified in a cell model after COVID-19 infection. This is a good positive control, indicating that our methodology is able to capture relevant biological processes. As expected, many pathways related with the immune system (adaptative [GO:0006954; FDR<1.23×10^-15^], innate [R-HSA-168249; FDR<5.03×10^-4^] and cytokine (GO:0034097;FDR<3.72×10^-14^]) were identified. More importantly, we identified several other pathways that can help to understand some of the reported long-term effects of COVID-19 infection, included brain-fog/dementia and cardiovascular problems.

Older adults, underserved populations, and persons with dementia are disproportionally impacted by the COVID-19 pandemic, showing a significantly increased infection risk as well as risk of severe illness from COVID-19 including neurocognitive consequences. In addition, several studies now suggest that COVID-19 can lead to Alzheimer’s-like dementia or other forms of neurocognitive impairment in persons free of cognitive impairment before infection, suggesting that COVID-19 can result in brain damage and neurocognitive impairment in individuals without preexisting neurological conditions. Our enrichment results for brain specific genes and AD related pathways (*38*) (Molecular Signatures Database M12921; FDR<5.46×10^-10^) strongly support CNS involvement in COVID-19 infection. Our results are in line with recent studies indicating a mechanistic overlap between AD and COVID-19 (*7*). Viral entry factors exist and are highly expressed in cells in the blood-brain barrier, potentially facilitating COVID-19 brain infection (*78*). Neuroinflammation is a major hallmark of AD, and COVID-19 (*10, 78*) infection has been shown to significantly alter known AD biomarkers implicated in brain inflammation. A recent study based on the expression of AD blood and CSF biomarkers in PBMCs and CSF of COVID-19 patients identified several protein biomarkers for AD, in line with a shared pathobiology of cognitive dysfunction in COVID-19 and AD. Our data further indicate that AD and general neurodegenerative biomarkers including GFAP, NFL and tau/MAPT proteins are altered in COVID-19 positive individuals and could also be used to identify individuals at risk of having long-term impact on memory loss, brain-fog and other brain-related outcomes. In the absence of effective treatments, a mechanistic understanding of how patients with AD become particularly vulnerable targets of SARS-CoV2, and how COVID-19 leads to cognitive impairment and AD in cognitively healthy individuals is critical to guide evidence-based healthcare management and development of targeted intervention. At this moment, our data cannot determine if individuals with altered levels of this AD-related proteins are more likely to develop AD or brain-fog and longitudinal studies are needed to confirm this hypothesis.

In addition to the CNS impairment, COVID-19 cases may also lead to an increased risk for CAD by increasing myocardial injury from viral infection (*79*). This is supported by the significantly enriched CAD related pathways and heart specific genes from our dysregulated proteins. Due to the complexity of the brain and heart, the damage of these two organs may be permanent and can cause post-acute COVID-19 syndrome (PACS) such as tiredness, dyspnea, chest pain, brain fog, and headache. Thus the dysregulated proteins belonging to the CAD related pathway (Cardiac Troponin T, ANGL4, FURIN, and AT1B1) might be used as prognostic tool to identify the cases at high risk for CAD and PACS in general, although additional follow-up studies are needed.

The majority of the identified 1,449 proteins are secondary cellular responses in different tissues due to viral infection and organ damage. We also aimed to identify proteins that are part of the causal mechanism for COVID-19 infection further associated with ventilation or death. By performing Mendelian randomization with the pQTLs of 1,437 proteins and GWAS for COVID- 19 infection in European population, we nominated BCAT2 and GOLM1 as causal proteins and increasing risk for COVID-19 infection. Together with the evidence that these two proteins were associated with COVID-19, agents that target BCAT2 and GOLM1 may be able to reduce the risk for both COVID-19 infection and COVID-19 related cognitive dysfunction

Our results using plasma pQTL as instrumental variables for MR analyses are significantly different to recent studies using eQTL. In a recent study, Gaziano (*80*) identified six genes (*CCR1, CCR5, IL10RB, IFNAR2, PDE4A* and *ACE2*) as causal repurposing candidates for COVID-19 using MR analyses with *cis*-eQTLs of 49 tissues in GTEx and the trans-ancestry meta-analysis of MVP and COVID-19 HGI. These discordant might be due to the lack of overlap between eQTLs and pQTLs. It is known that mRNA levels do not necessarily correlate well with protein levels (*46*). Our previous study demonstrated a low level of overlap between expression and protein QTLs (*46*). In fact, in the large pQTL atlas used for MR in this study, no cis-pQTL was found for those proteins. As protein are closer to traits that expression levels, eQTL can lead to many false positive and negative findings. The limitation of our MR analyses is the causal inference of BCAT2 and GOLM1 for COVID-19 infection cannot be generalized to other ethnic groups due to lacking pQTLs resources for African and other ethnic groups. Further investigation of the roles of these two proteins in other ethnic groups is needed. MR is also limited to those proteins that have a strong pQTL, therefore this approach might miss many potential therapeutic targets. For this reason, we used additional Bayesian approach to identify key-drivers in the proteins network that could be also part of the causal pathway and potentially druggable. Gene network analyses identified 141 key proteins, of which 35 have known drug targets, some of them already being tested in several clinical trials. Additional studies will be needed to confirm that the proposed targets and FDA compounds have an impact on COVID-19 outcomes.

In summary, we performed deep proteomics profiling of COVID-19 patients, identifying dysregulated proteins associated with severe outcomes. We created a web portal (https://covid.proteomics.wustl.edu/) for users to interactively explore these plasma proteomic signatures of COVID-19 infection and outcomes. We created prediction models for ventilation and death, presented AD and CAD related changes with COVID-19 infection, and identified two novel potential druggable targets. Our findings show the promise of proteomic studies contributing to the understanding of COVID-19 biology and pathophysiology.

## Materials and Methods

### Sample Collection

We recruited 332 COVID-19 patients with positive SARS-CoV-2 test from March 26, 2020 to September 30, 2020 at Barnes Jewish Hospital, St. Louis Children’s Hospital or affiliated Barnes Jewish Hospital testing sites in St. Louis. We obtained the ventilation status and death status from the hospital records. Using plasma samples collected prior to November 2019, we selected 150 controls by matching on sex, race and age. We obtained the informed consent for enrollment in the study from 482 participants. The study was approved by institution review board at Washington University School of Medicine in St. Louis.

### Plasma Proteomic data

Peripheral blood was collected, and plasma was isolated by centrifuge and stored at -80⁰C. The proteomic data in plasma was measured using SomaScan v4.1 7K, a multiplexed, single-stranded DNA aptamer-based platform (*81*) from SomaLogic (Boulder, CO). Three different dilution sets (1:5, 1:200, 1:20,000) were used for plasma samples. Instead of physical units, the readout in relative fluorescent units (RFU) was used to report the protein concentration targeted by 7,584 modified aptamers. To mitigate technical variation introduced by the readout, pipetting errors, inherent sample variation, initial data standardization at the sample and aptamer level including hybridization control normalization, median signal normalization and inter-plate calibration was performed by SomaLogic with control aptamers (positive and negative controls) and calibrator samples (*82*). Hybridization control normalization on each plate was used to correct for systematic variability in hybridization. Median signal normalization was performed within the same dilution group to remove the bias due to pipetting variation, variation in reagent concentrations, and assay timing among others. Finally inter-plate Calibration based on calibrator samples was performed separately on each aptamer to remove plate bias.

We further processed the proteomics data using SomaDataIO (v1.8.0) (*81*) and Biobase (v2.42.0) (*83*) and removed the aptamer outliers and sample outliers by using the following approaches (*46*) (Fig. S2): 1) Aptamer outliers had greater than 15% samples with concentration below 2 standard deviation above the average RFU level of the dilution buffer. 2) Aptamer outliers had greater than 0.5 of maximum difference between aptamer calibration factor and median of plate calibration factor. 3) Aptamer outliers had median coefficient of variation (CV) more than 0.15. 4) Aptamer outliers had greater than 15% of samples whose log10 transformed RFU level fell outside of either end of 1.5 fold of Interquantile range (IQR). 5) Samples outliers had more than 15% aptamers falling outside of either end of 1.5 fold of IQR . 6) Aptamer outliers shared by ∼80% of sample outliers.

All 7,584 aptamers were annotated with Universal Protein Resource (UniProt)(*84*) identifiers and Entrez Gene symbols. After QC, 7,055 aptamers and 482 individuals remained and were used for differential abundance analyses.

### Statistical Analyses

To identify the plasma biomarkers associated with COVID-19 infection and severity in the discovery cohort, protein levels were first log10 transformed to approximate the normal distribution. Regression analysis was then performed using lm function in R (R version 3.5.2 (2018-12-20)) with:

log10 (protein abundance level) = β0 + β1 * outcome + β2 * age + β3 * gender + β4 *ethnicity + β5 * plate

where an outcome corresponds to each of three COVID-19 outcomes (infection, ventilation and death) and plate denotes 8 different plates used by SomaScan. Volcano plots were created with the ggplot2 package and Venn diagrams were created with R VennDiagram package.

The proteomic data for 482 individuals were assayed in 8 different plates. Even though the intraplate and interplate variation of aptamer RFU levels has been normalized and the plate was used as covariate in our analyses to control for batch effects, there may be unobserved hidden factors which correlate with the outcomes and biased our estimate. Thus, we estimated these hidden factors from the matrix of 7,055 proteins for 482 individuals using surrogate variable analysis (SVA) implemented in R sva package. Since sva estimation requires no missing data, we first imputed the missing data points by random sampling of non-missing data points for each protein separately by COVID-19 cases and controls. Then number of surrogate variables was estimated using a permutation procedure originally proposed by Buja and Eyuboglu(*85*) (“BE” method) and asymptotic approach proposed by Leek(*86*) (“LEEK” method). BE method yielded 29 surrogate variables and LEEK method yielded 3 surrogate variables for our proteomic data. Regression analysis was performed using lm function in R for 3 outcomes with 1) log10 (protein abundance level) = outcome + age + gender + ethnicity + 29 BE surrogate variables; 2) log10 (protein abundance level) = outcome + age + gender + ethnicity + 3 LEEK surrogate variables. Finally we compared the effect estimates from these two regression models with the main model shown above and calculated the Pearson correlation coefficient of the effect size among the three models (Fig. S3).

### Replication Strategy

To replicate the proteins associated with COVID-19 infection and severity, we searched publicly available COVID-19 resource and downloaded plasma proteomic data from MGH COVID-19 cohort (*23*). They recruited 384 individuals (306 cases and 78 controls) and assayed 5,284 aptamers using SomaScan at the first time point. As this proteomic data included neither calibrators nor negative/positive controls, we performed only 2 rounds of IQR strategy to identify the aptamer outliers and individual outliers. After QC, 373 individuals (297 cases and 76 controls) and 5,258 aptamers remained. Among these aptamers, 4,301 proteins were present in both the discovery and replication data and were used for replication analyses using the following model:

log10 (protein abundance level) = β0 + β1 * outcome + β2 * age-range

Gender, race and plate information were not available in this public data and were not included in the covariates. In addition, instead of actual age, age range (1 = 20-34, 2 = 36-49, 3 = 50-64, 4 = 65-79 and 5 = 80+) was provided. We used COVID-19 (“COVID” column), ventilation (ventilated within 28 days) and death (death within 28 days) as each of three outcomes.

### Meta-analyses

For 4,301 proteins present in both discovery and replication cohort, we performed meta-analysis to identify proteins significant in both discovery and replication samples and boost the statistical power of our analyses. We meta-analyzed the differential abundance analysis results from discovery and replication cohorts using the inverse variance weighed fixed effect model (“SCHEME STDERR”) implemented in METAL (*87*). The Bonferroni corrected threshold (1.16×10^-5^) was used in the meta-analysis to correct for multiple testing.

### Prediction Model

Least absolute shrinkage and selection operator (Lasso) regression performs both variable selection and regularization, which forces the coefficients of several weak variables as zero and excludes them in the model. For each of the 3 outcomes, we used five-fold cross validation and Lasso regression in caret R package and selected important features from proteins associated with COVID-19 (n=841), ventilation (n=833) and death (n=253). Besides these 3 sets of protein, we also evaluated the performance of prediction model for COVID-19 and severity using the proteins consistent across all 3 outcomes.

In addition, to construct a ventilation specific prediction model, we selected 50 proteins using the following criteria: 1) they were significant in discovery and replication for ventilation (FDR<0.05), 2) significant in the combined (discovery and replication) meta-analysis at Bonferonni corrected threshold (P < 0.05/4,301=1.16×10^-5^) for ventilation, 3) concordant direction of effects between discovery and replication for ventilation, and 4) not significant in the meta-analysis for death (P>0.05). From these proteins, Lasso regression further selected 32 and evaluated as a separate prediction model for ventilation.

Similarly, for death specific prediction model, 5 death specific proteins (ASB9, Endocan, jun-D, EGFLA, and Glutathione S-transferase Pi) were selected using the following criteria:1) they were significant in discovery and replication for death (FDR<0.05), 2) significant in the combined (discovery and replication) meta-analysis at Bonferroni corrected threshold (P < 1.16×10^-5^) for death, 3) concordant direction of effects between discovery and replication for death, and 4) not significant in the meta-analysis for ventilation (P>0.05).

ROC curve and AUC statistics were generated using pROC package. To evaluate the performance of these proteins, the positive predictive value (PPV) and negative predictive value (NPV) were estimated based on Youden’s J statistic optimal cut-off using pROC coords function. The COVID-19 samples who were ventilated and died (44 in discovery and 21 in replication) were removed in the evaluation process. For ventilation and death prediction, both discovery and replication samples were unbalanced and included more controls than cases. To compute NPV and PPV, we first selected the age, gender and ancestry matched controls for each of ventilated cases and each of died cases separately in discovery cohort and age matched controls for each of ventilated cases and each of died cases separately in replication cohort. Then bootstrap sampling of these case-control balanced subsamples was repeated 100 times separately for ventilation and death to increase the estimation accuracy. The PPV and NPV was calculated based on the average of PPV and NPV across 100 bootstrap samples.

### Pathway Enrichment Analyses and Tissue Specificity Enrichment Analyses

Using default background gene lists, we performed pathway enrichment analyses using Enrichr (*25*) and FUMA GWAS (*24*) separately for proteins associated with COVID-19 infection (n=841), ventilation (n=833), death (n=253) and 64 proteins consistent across 3 outcomes. Both of these approaches applied over-representation analyses by calculating the amount of input genes that belong to the certain pathways using Fisher’s exact test. Pathways of “COVID-19 related gene sets 2021” were tested for the significant proteins using Enrichr. Gene2Func was used in FUMA and the gene sets of pathways included multiple databases including GO biological processes, Reactome gene sets, KEGG pathways, canonical pathways, PANTHER pathways and Wiki pathways. Using the same 4 protein sets, tissue specificity enrichment test based on GTEx v8 tissue types was also performed using Gene2Func in FUMA. The significance was defined as Benjamini-Hochberg FDR<0.05. The ggplot2 R package was used to create the dot chart and tile plots for the significant enriched pathways and significant enriched tissues.

### Mendelian randomization (MR) analyses

We downloaded the COVID-19 GWAS results by COVID-19 host genetics initiative for COVID-19 vs. population (COVID19_HGI_C2_ALL_eur_leave_23andme_20210107.txt.gz) and GWAS results of 4,907 plasma proteins in 35,559 Icelanders (*60*). Since genetic variants were randomly inherited from parents at birth which validate their occurrence before any biological traits and diseases, this has been applied in Mendelian randomization (MR) analyses. MR uses genetic variants of exposures as a natural experiment to investigate the causal relations between modifiable exposures and disease. Two-sample MR implemented in R package TwoSampleMR (*88*) version 0.5.5 was performed to identify the causal plasma biomarkers using each of 1,437 differential abundant proteins identified by our analyses with GWAS results as exposure and COVID-19 infection as outcome. First, non-palindromic SNPs with association P < 1.5×10^-6^ for each assessed protein were selected and independent genetic variants was identified using clump_kb=10000 and clump_r2=0.001. Then these independent variants were extracted from outcome (COVID-19 infection) GWAS results. Before MR analyses, data harmonization step including fixing the strands, and variants with non-matching alleles were removed. One critical assumption for TwoSampleMR is no horizontal pleiotropy. We performed MR pleiotropy residual sum and outlier (MR-PRESSO) and directional pleiotropic effects by egger regression intercept test (MR-Egger) to check and correct for possible pleiotropy. First, if global horizontal pleiotropy was confirmed by MR-PRESSO, the outlier SNPs were removed from the instrument lists and corrected P was selected from MR-PRESSO. Second, if there was no evidence of global horizontal pleiotropy and there was significant egger intercept, MR-Egger P was used. Third, if both global horizontal pleiotropy and egger intercept were not significant, inverse variants weighted meta-analysis (IVW), aggregating all of single-SNP causal effects, was used. The Bonferroni corrected threshold (0.05/1437=3.48x10^-5^) was used as statistical significance for MR estimates..

### Co-localization analyses

We performed Bayesian colocalization analyses using coloc.abf function in coloc R package (version 5.1.1) (*89*). Using the default priors as P1=1×10^-4^, P2=1×10^-4^ and P12=1×10^-5^, the posterior probability of co-localization was estimated with PP.H4 > 0.8 which indicates both proteins and disease shared the same causal variants.

### Hub and druggable proteins

To identify proteins potentially driving infection or severity, we utilized multi-scale embedded gene co-expression network analysis (MEGENA), a co-expression network analysis technique, to identify the most connected “hub” proteins (*62*). Because these proteins share connections with many others, they are predicted to contribute to disease status or severity. The MEGENA pipeline was used to analyze a set combining the differentially abundant proteins from all three analyses (1, 449). Proteins with both increased and decreased abundance were included for each set. A matrix of protein measurements and samples was used as the input parameter. MEGENA consists of three main steps; first, a fast planar filtered network is constructed. To do this, MEGENA first calculates abundance similarity between each protein pair. We utilized Pearson correlation to measure similarity. We performed false-discovery rate filtering of the protein abundance similarities at a p-value threshold of 0.05, done by calculating pairwise p-values with Fisher’s Z-transformation. We computed correlations based on protein levels from COVID-19 positive individuals only. To build the planar filtered network of protein connections, MEGENA starts from a network of protein nodes with no connections. It then iteratively orders protein pairs by their co-expression and tests for planarity. If the pair passes planarity, an edge between the proteins is added. This process is repeated until the number of edges is equal to three times number of vertices -2).

Next, MEGENA performs multiscale clustering analysis on the finalized planar filtered network. This is done through optimization of the local clustering structure through local path index. Because MEGENA calculates clusters and identifies hubs using a permutation-based approach, we performed the analysis ten times for each of the four sets of proteins and pooled the hub proteins identified by each run. This produced four lists of hub proteins, each identified in one of the sets of proteins. Each of the ten runs performed 100 permutations. Clusters were limited to a size of number of nodes/2 on the high end and ten on the low end. Bonferroni-corrected P < 0.05 is used to identify hub proteins.

For each of the hub proteins, we characterized them based on their presence in enriched pathways identified through our earlier analysis, including pathways associated with COVID-19, immune response, and cell surface proteins among others. Additionally, each protein name was used as a query into UniProt (*84*), which provides information on drugs associated with each protein curated by DrugBank (*90*). These drugs may be useful for repurposing for COVID-19 treatment or prevention. To visualize the correlations between the proteins for each analysis, we utilized the RCy3 (*91*) R package to export the igraph object produced by MEGENA into Cytoscape (*92*). Using RCy3, we were able to customize the appearance of each of the nodes based on their status as a hub protein, the cluster they were a part of, and the number of significant correlations for each protein.

We also performed MEGENA analysis using a matrix consisting only of the hub proteins identified from the combined analysis. This was performed only to identify clustering of the hub proteins; we did not investigate the “hubs of the hubs”, although by default MEGENA identifies them. We used the same criteria for identification of clusters, with a minimum of ten nodes per cluster and a maximum of n/2. This analysis was performed once with 100 permutations. We customized nodes by their association with drugs in DrugBank, their identification as hub proteins in each of the four analyses, and their classification in each cluster identified by MEGENA.

We produced a network plot for each of the four analyses and for the hub proteins specifically identified in the combined analysis of all differentially abundant proteins. Using RCy3, we imported the igraph object produced by MEGENA based on the correlation calculations. We customized the appearance of each of the nodes based on their status as a hub protein, the cluster they were a part of, and the number of significant correlations for each protein. First, the lowest- level cluster identified by MEGENA determines the color of each node/protein. For example, a protein may be part of a large cluster three, as well as subclusters 25 and 79. As cluster 79 is the most specific cluster, the protein is colored corresponding to that cluster. These colors were manually matched to every cluster by supplying random RGB color codes. Node size was changed based on the number of edges containing that node, with larger nodes corresponding to higher connectedness. Finally, node shape was determined by hub protein status; diamonds represent hubs, while non-hubs are circular.

## Data Availability

All data produced are available online at https://www.niagads.org/Knight ADRC-collection

https://covid.proteomics.wustl.edu/

## Acknowledgements

We thank all the participants and their families, as well as the many involved institutions and their staff.

We acknowledge SomaLogic Operating Co., Inc. as the provider of the proteomic data measured using the modified aptamer-based SomaScan® Assay. SomaScan®, SOMAmer® and SomaSignal™ are trademarks of SomaLogic Operating Co., Inc.

This work was supported by access to equipment made possible by the Hope Center for Neurological Disorders, the Neurogenomics and Informatics Center (NGI: https://neurogenomics.wustl.edu/) and the Departments of Neurology and Psychiatry at Washington University School of Medicine.

## Author contributions

Conceptualization: CC YJS, Methodology: CC, YJS, LW BZ, WS, and DW performed the analyses; Supervision: CC, YJS; Data generation and QC: JT, CR, JN, JB, OHB, JO, RP, CWR, PAM, Funding: SC, CC, DJ, MG, ART, SC, Writing – original draft: LW, DW, YJS, CC; Writing – review & editing: All authors reviewed and approved the manuscript.

## Data and material availability

Proteomic data from the Knight ADRC participants are available at the NIAGADS and can be accessed at https://www.niagads.org/Knight <u>ADRC-collection</u>;

The summary results using these data are also available to the scientific community through a public web browser: https://covid.proteomics.wustl.edu/

## Supplementary Materials

The PDF file includes:

Figs. S1 to S13

**Other Supplementary Materials for this manuscript included the following:**

Tables S1 to S22

---

**Fig. S1.**
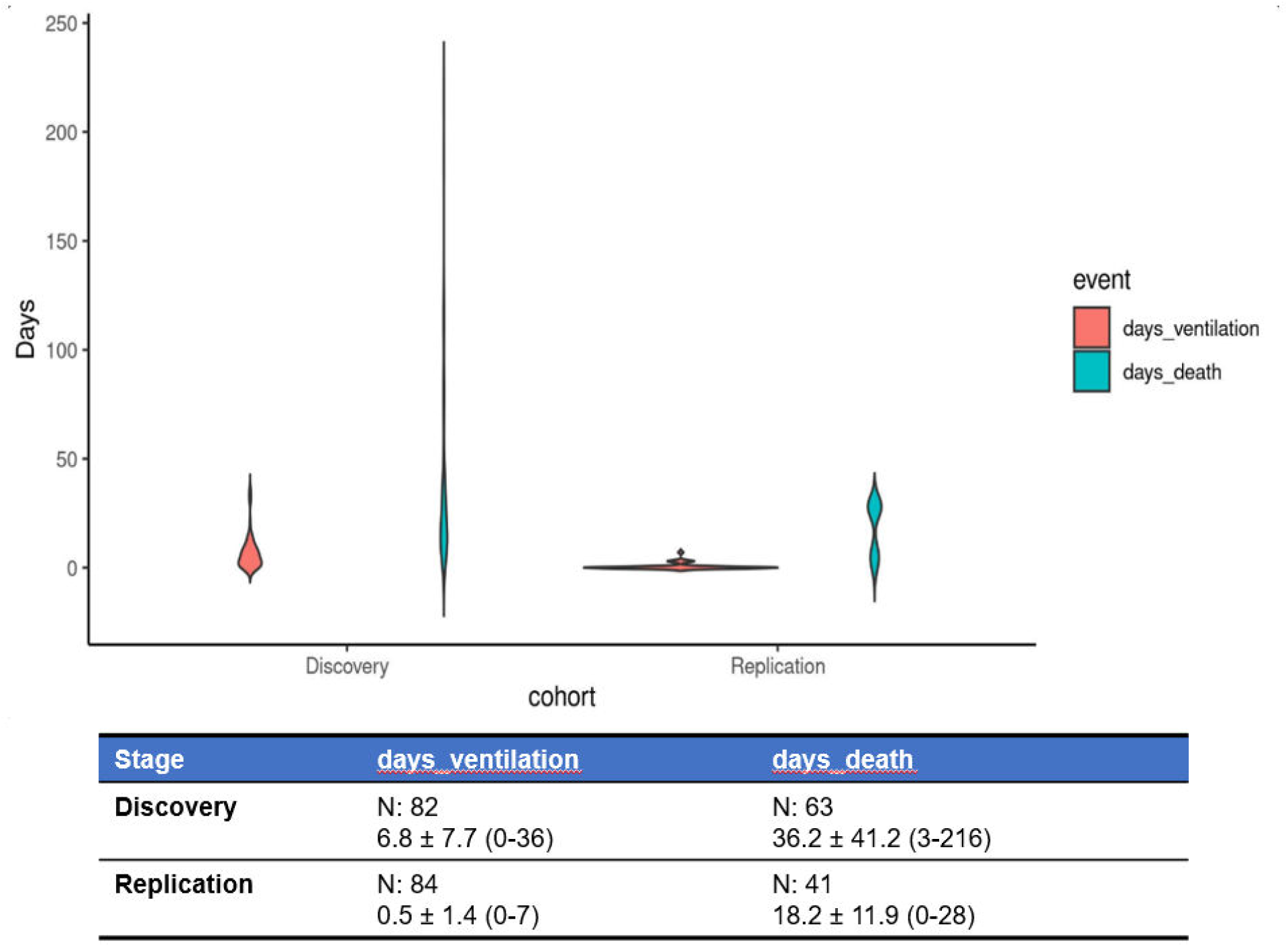
Violin plots of number of days to ventilation and death in discovery and replication. The table of sample size (N) and mean and standard deviation (mean ± SD) for number of days to ventilation and number of days to death.

**Fig. S2.**
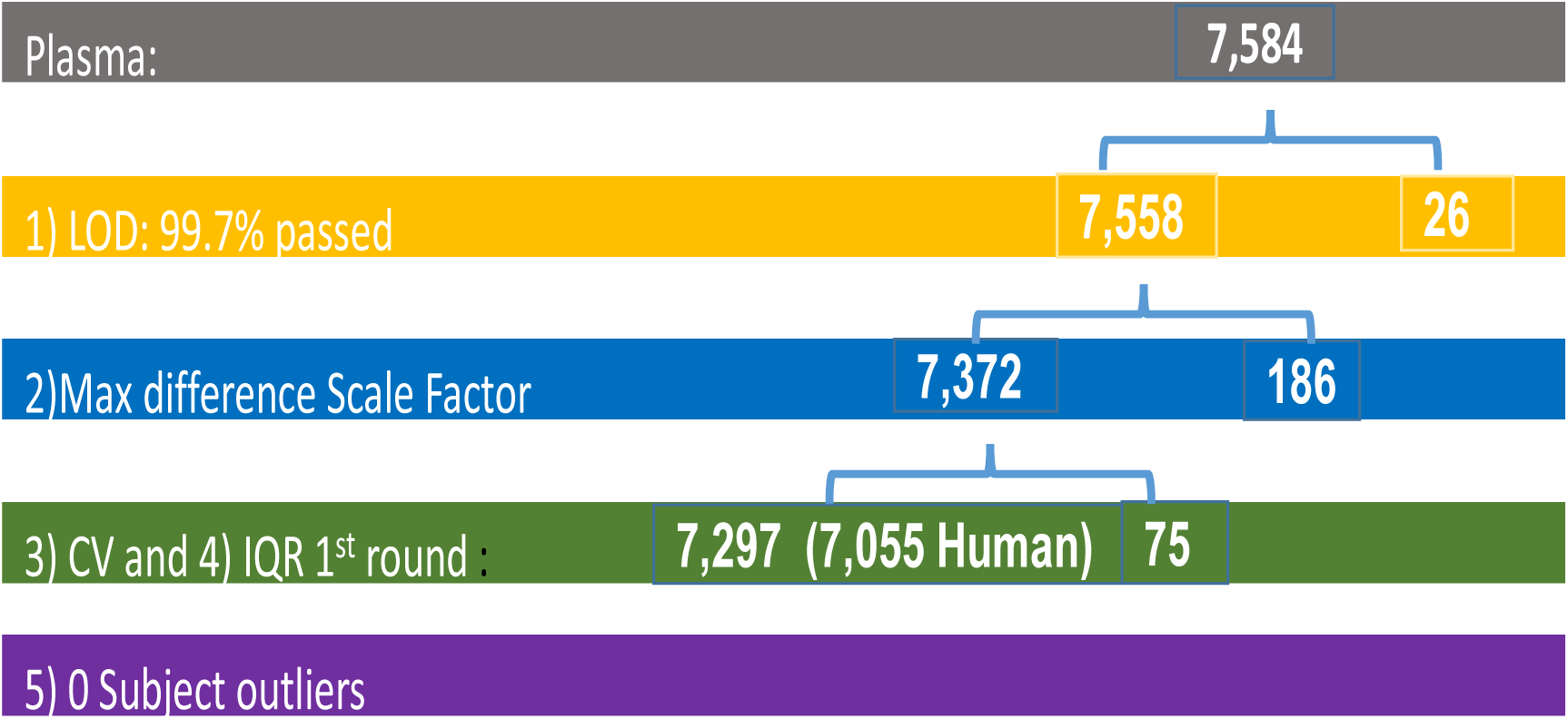
Quality control flow for plasma SomaScan v4.1 7K data in discovery stage. The number of aptamer kept (left side number) and removed (right side number) in each step was include in this figure. In summary, 7,297 (92.8%) aptamers were kept. Among the 7,297 aptamers, 7,055 which targets human proteins were used in our analyses and 4,301 were available in replication data. LOD represents limit of detection. CV denotes coefficient of variation. The detailed explanation of quality control for each step can be found in Materials and Methods.

**Fig. S3.**
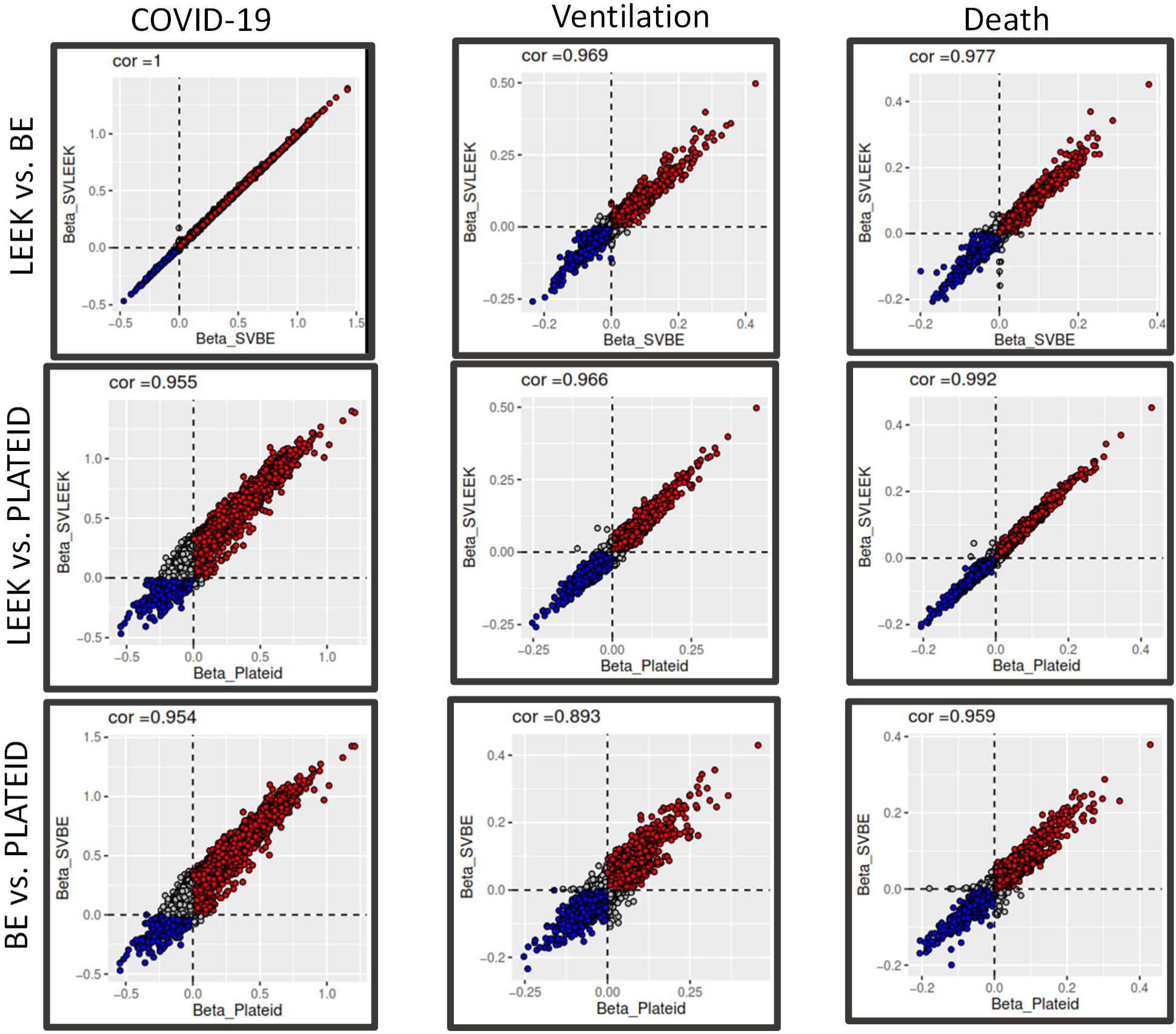
Scatterplot of effect size for including 3 “LEEK” method estimated surrogate variables, 29 “BE” method estimated surrogate variables and Plateid as covariates in the model. The surrogate variables were estimated using 7,055 proteins assayed for 482 samples using R package sva. Then linear regression was performed including 3 “LEEK” method estimated surrogate variables, or 29 “BE” method estimated surrogate variables, or plate (PLATEID) as covariates to estimate the effect size for 3 outcomes. The effect size from the 3 approaches were similar with very high Pearson correlation coefficient among them.

**Fig. S4.**
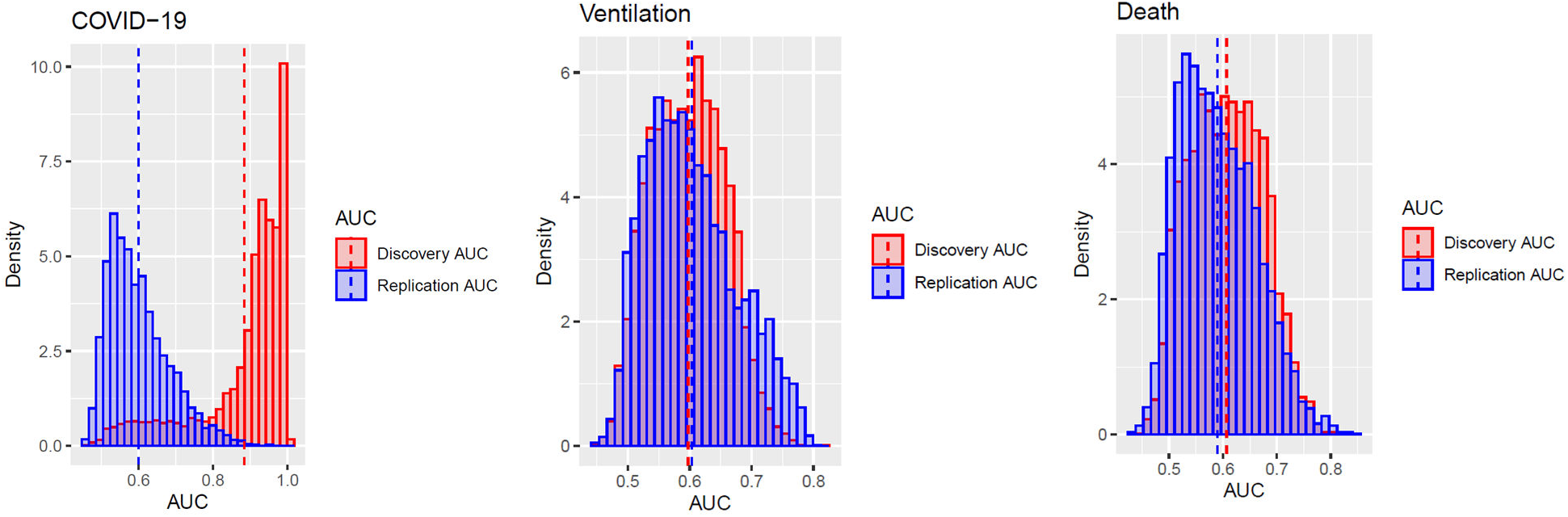
Distribution of AUC for COVID-19 infection, Ventilation status and Death status. AUC was estimated separately by discovery and replication stage from univariate logistic regression model using each of COVID-19 infection, Ventilation status and Death status as the response variable, and each of 4301 proteins as the predictor. The histograms of AUC (AUC displayed along x-axis and Density displayed along y-axis) were plotted separately for COVID-19 infection, Ventilation status and Death status using ggplot2 R package (red color: AUC in discovery stage; blue color: AUC in replication stage).

**Fig. S5.**
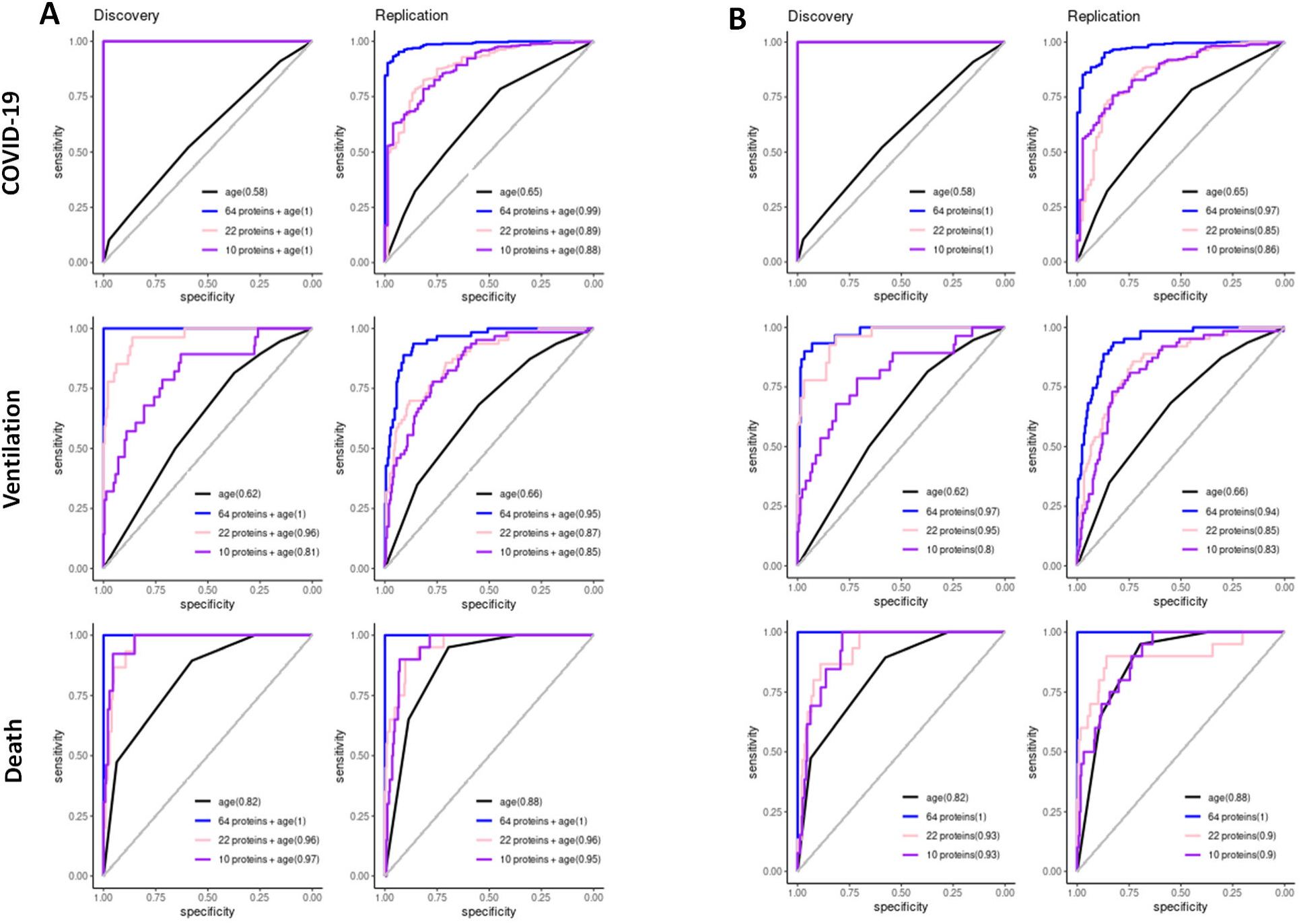
The ROC curves using 64 significant proteins consistent across all 3 outcomes, 22 proteins selected by Lasso regression for ventilation, and 10 proteins selected by Lasso regression for death. A) The ROC curves with corresponding AUC in each of discovery and replication stage derived from logistic regression model (univariate or multivariate) using each of COVID-19, ventilation and death as response variables, either age along (black solid curves) as predictor, or age together with 64 ventilation proteins (blue solid curves) as predictors, or age together with 22 proteins (pink solid curves) as predictors, or age together with 10 proteins (purple solid curves) as predictors. Y-axis represents sensitivity and x-axis represents specificity. All these 3 sets of proteins demonstrated significantly higher AUCs (0.81-1) for ventilation and death than the model using age alone. **B)** The ROC curves with corresponding AUC in each of discovery and replication stage derived from logistic regression model (univariate or multivariate) using each of COVID-19, ventilation and death as response variables, either age along (black solid curves) as predictor, or 64 ventilation proteins (blue solid curves) as predictors, or 22 proteins (pink solid curves) as predictors, or 10 proteins (purple solid curves) as predictors. Y-axis represents sensitivity and x-axis represents specificity. All these 3 protein set also demonstrated significantly higher AUCs (0.8-1) for ventilation and death than the model using age.

**Fig. S6.**
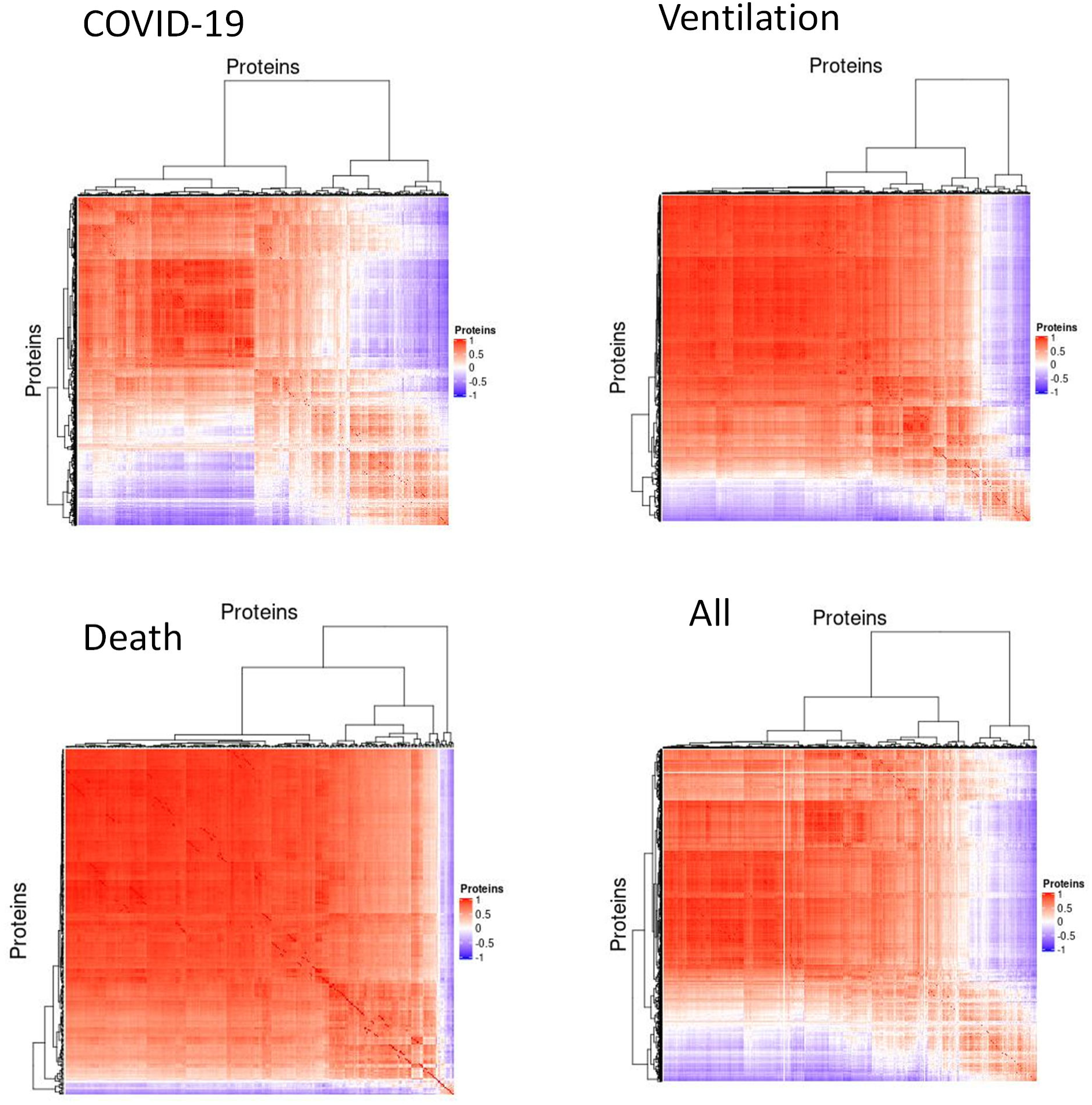
Heatmap of Pearson correlation matrix of detected and replicated significant proteins for COVID-19, Ventilation, Death and all 3 outcomes combined. Pearson correlation matrix for the significant differential abundant proteins (841 for COVID-19, 831 for ventilation, 253 for death and 1449 for all 3 outcomes combined). ComplexHeatmap R package was used to plot the heatmap.

**Fig. S7.**
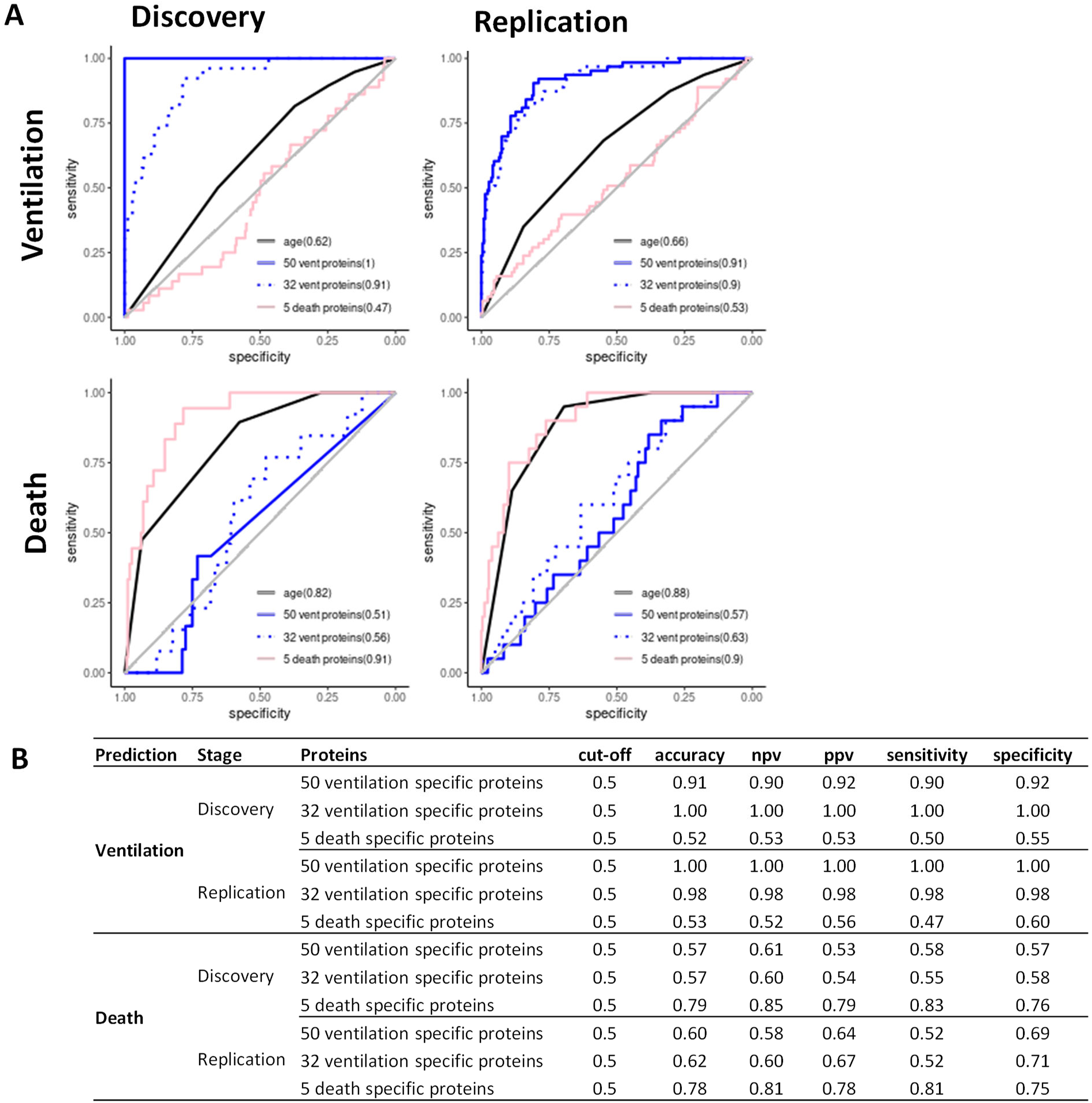
Performance of prediction model using ventilation specific and death specific proteins. A) The ROC curves with corresponding AUC in each of discovery and replication stage derived from logistic regression model (univariate or multivariate) using each of ventilation and death as response variables, either age along (black solid curves) as predictor, or 50 ventilation specific proteins (blue solid curves) as predictors, or 32 Lasso selected ventilation specific proteins (blue dotted curves) as predictors, or 5 death specific proteins (pink solid curves) as predictors. Y-axis represents sensitivity and x-axis represents specificity**. B)** The table of accuracy, negative predictive value.(NPV), positive prediction value (PPV), sensitivity and specificity at 0.5 of Youden’s J statistic for the logistic regression model used in b. This evaluation used the case control balanced subsample by selecting age and gender matched or age matched controls in discovery and replication respectively for each of ventilated cases or died cases. As expected, for ventilation prediction, both 50 and 32 ventilation specific proteins demonstrated ∼90% -100% of these evaluations. Whereas for death, 5 death specific proteins showed ∼75%-85% of these evaluations.

**Fig. S8.**
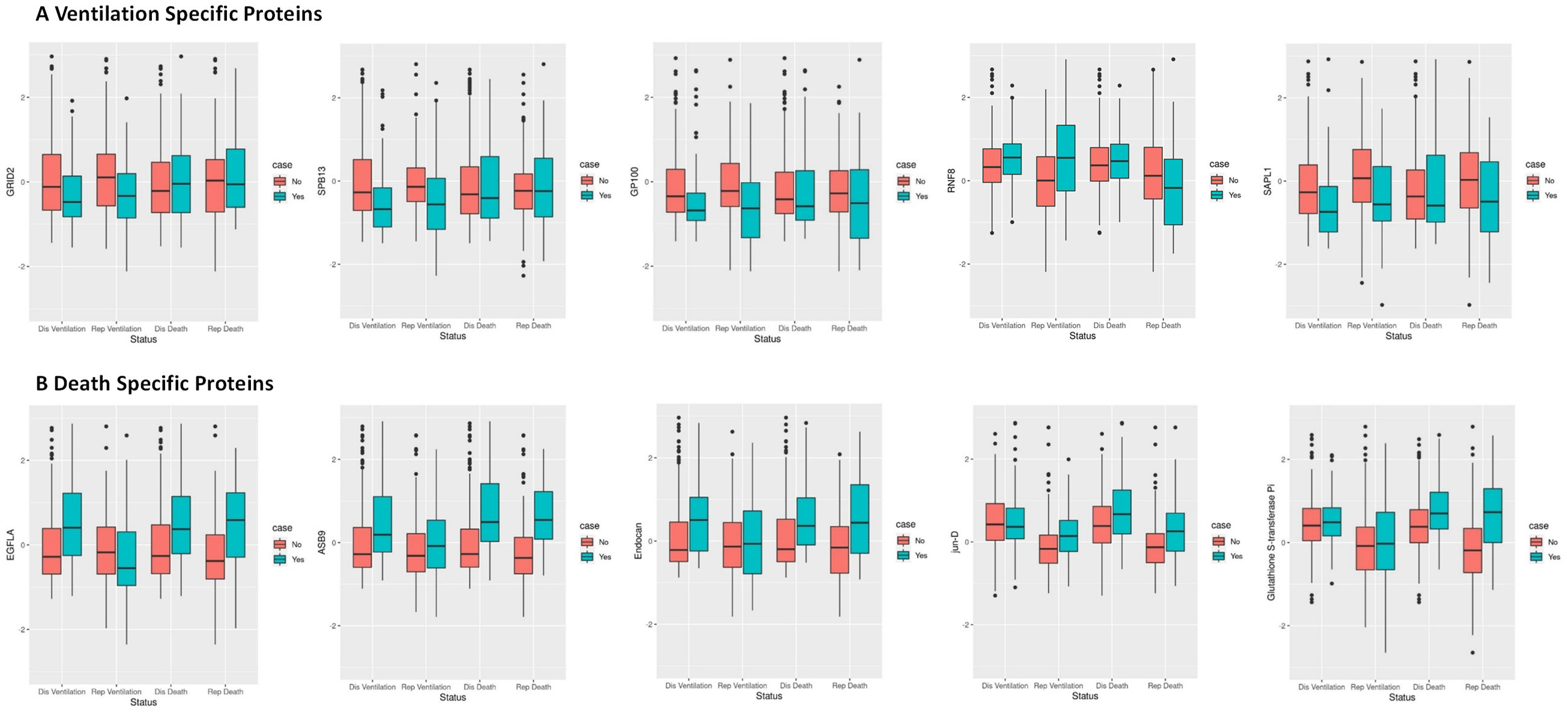
Boxplots of Ventilation specific proteins and Death specific proteins. A) Boxplots of 5 ventilation specific proteins (GRID2, SPB13, GP100, RNF8, SAPL1) by Status (Dis Ventilation denotes Ventilation status in Discovery; Rep Ventilation represents Ventilation status in Replication; Dis Death denotes Death status in Discovery; Rep Death represents Death status in replication). The y-axis was Z-Score of protein levels which converted separately in discovery stage and replication stage. Horizontal bars demonstrate the median value. The lower and upper borders of the box represent the first and the third quantitle. Dis: Discovery; Rep: Replication**. B)** Boxplots of 5 death specific proteins (EGFLA, ASB9, Endocan, jun-D, Glutathione S-transferase Pi) by Status (Dis Ventilation denotes Ventilation status in Discovery; Rep Ventilation represents Ventilation status in Replication; Dis Death denotes Death status in Discovery; Rep Death represents Death status in replication). The y-axis was Z-Score of protein levels which converted separately in discovery stage and replication stage. Horizontal bars demonstrate the median value. The lower and upper borders of the box represent the first and the third quantitle. Dis: Discovery; Rep: Replication

**Fig. S9.**
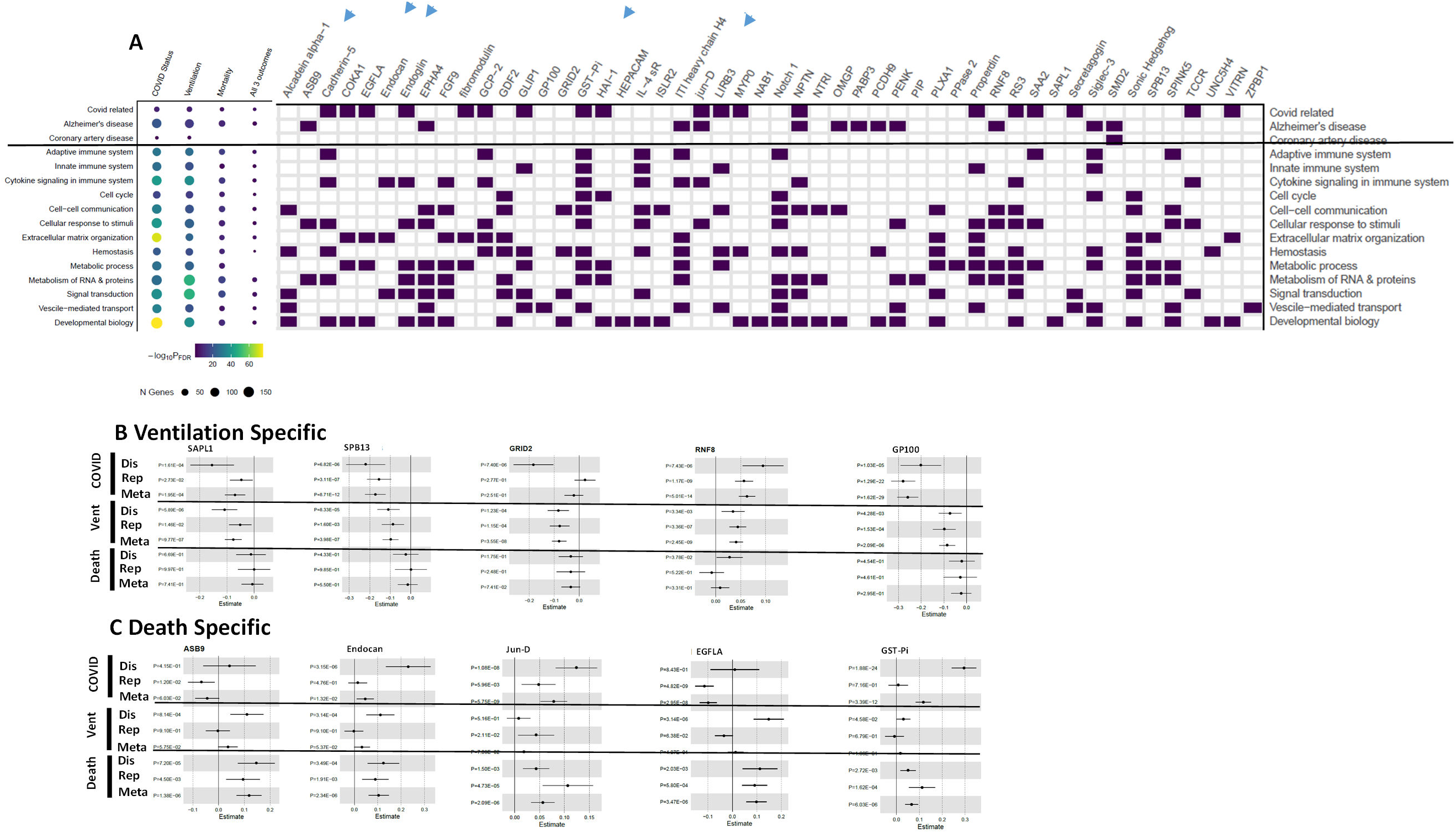
Pathway enrichment analyses using the Entrez Gene Symbol of the identified significant proteins (45 ventilation specific proteins and 5 death specific proteins included in the figure). A) Entrez gene symbol of the identified significant proteins were used in pathway enrichment analyses via Enrichr and FUMA. 45 ventilation specific proteins and 5 death specific proteins were enriched for 16 pathways. 19 of these proteins were enriched in covid related pathways, 12 of these proteins were enriched in Alzheimer’s disease. And one protein SMD2 (*SNRPD2*) was involved in Coronary artery disease. Blue arrows point to the death specific proteins. **B)** Forest plots of ventilation specific proteins: Proactivator polypeptide-like 1 (SAPL1), Serpin B13 (SPB13), Glutamate receptor ionotropic, delta-2 (GRID2), E3 ubiquitin-protein ligase RNF8 (RNF8), Melanocyte protein PMEL (GP100**) C)** Forest plots of death specific proteins: Ankyrin repeat and SOCS box protein 9 (ASB9), Endothelial cell-specific molecule 1 (Endocan), Transcription factor JunD (jun-D), Pikachurin (EGFLA), Glutathione S-transferase Pi (GST-Pi).

**Fig. S10.**
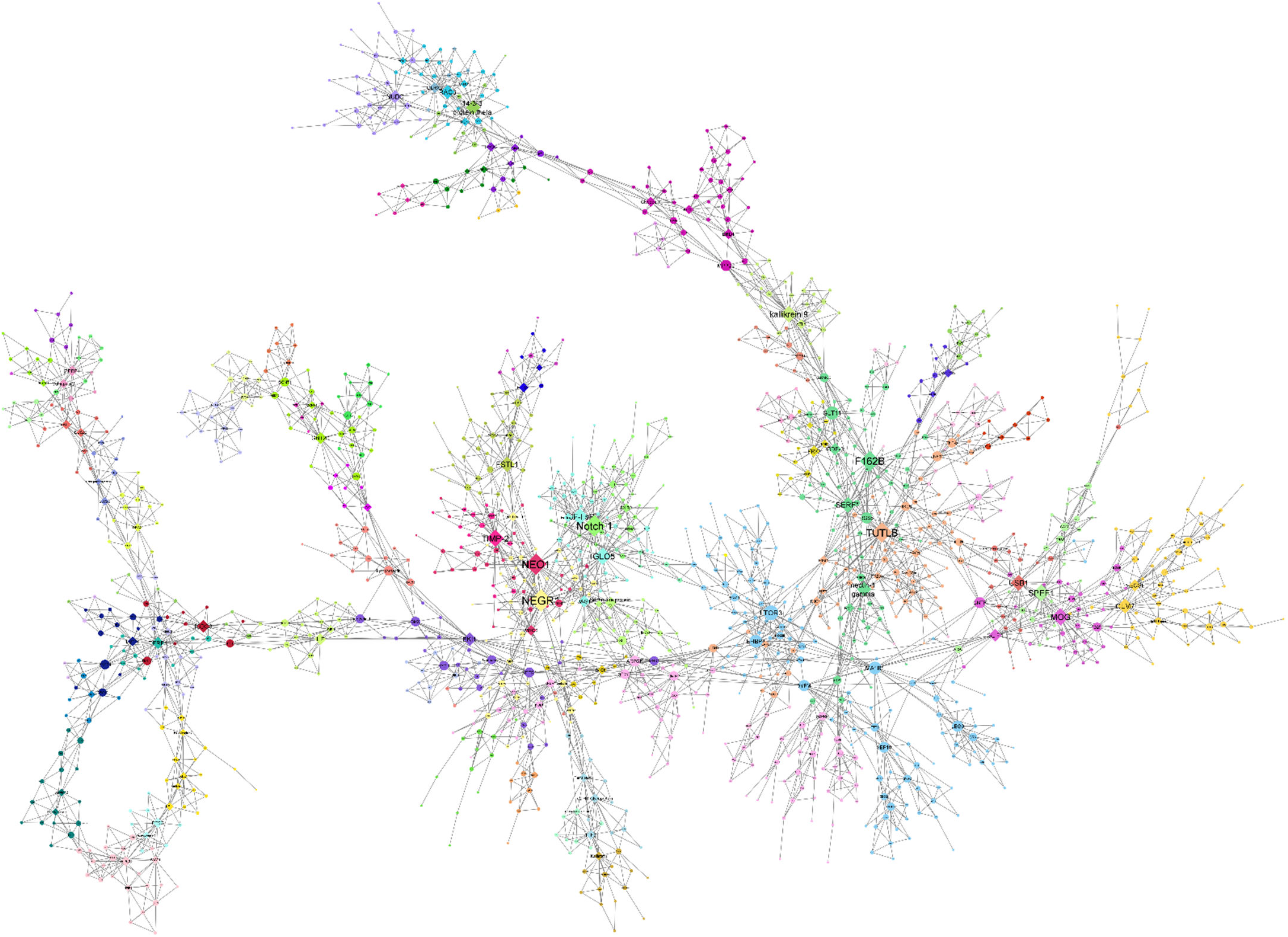
MEGENA network plot of 1,449 differentially abundant proteins in any of three analyses. MEGENA network plot produced using the igraph object produced by MEGENA analysis of proteins differentially expressed in any of case vs control status, ventilation status, or death status. Colors represent clusters of proteins identified by MEGENA. Shape corresponds to hub protein status; diamonds represent hubs, all other proteins are circular. Node size corresponds to the connectedness of that protein as determined by significant correlations, scaled by number of edges connecting that node.

**Fig. S11.**
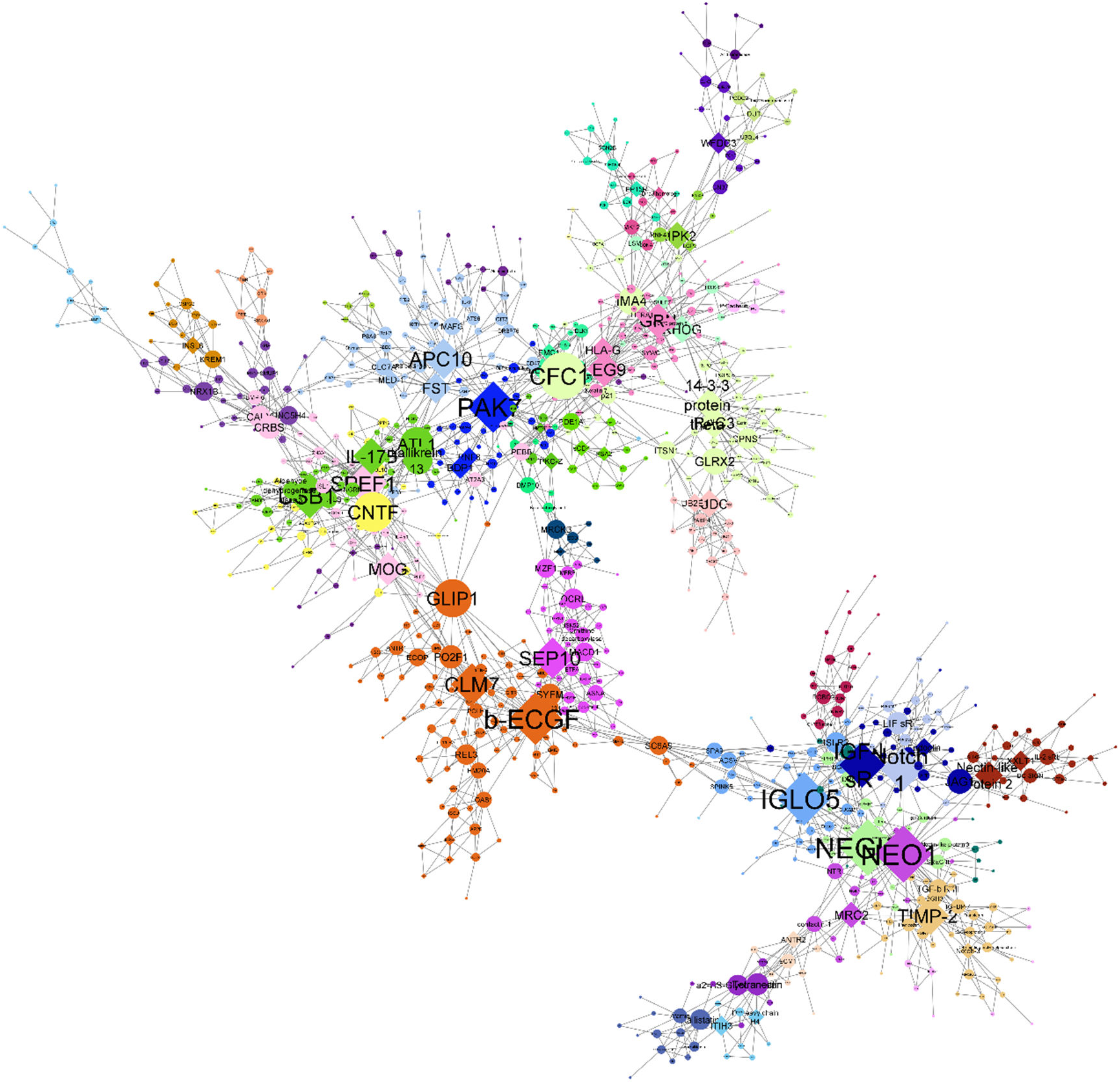
MEGENA network plot of 841 proteins differentially abundant in COVID-19 case vs control status. MEGENA network plot produced using the igraph object produced by MEGENA analysis of 841 proteins differentially expressed in COVID-19-positive individuals vs unaffected controls. Colors represent clusters of proteins identified by MEGENA. Shape corresponds to hub protein status; diamonds represent hubs, all other proteins are circular. Node size corresponds to the connectedness of that protein as determined by significant correlations, scaled by number of edges connecting that node.

**Fig. S12.**
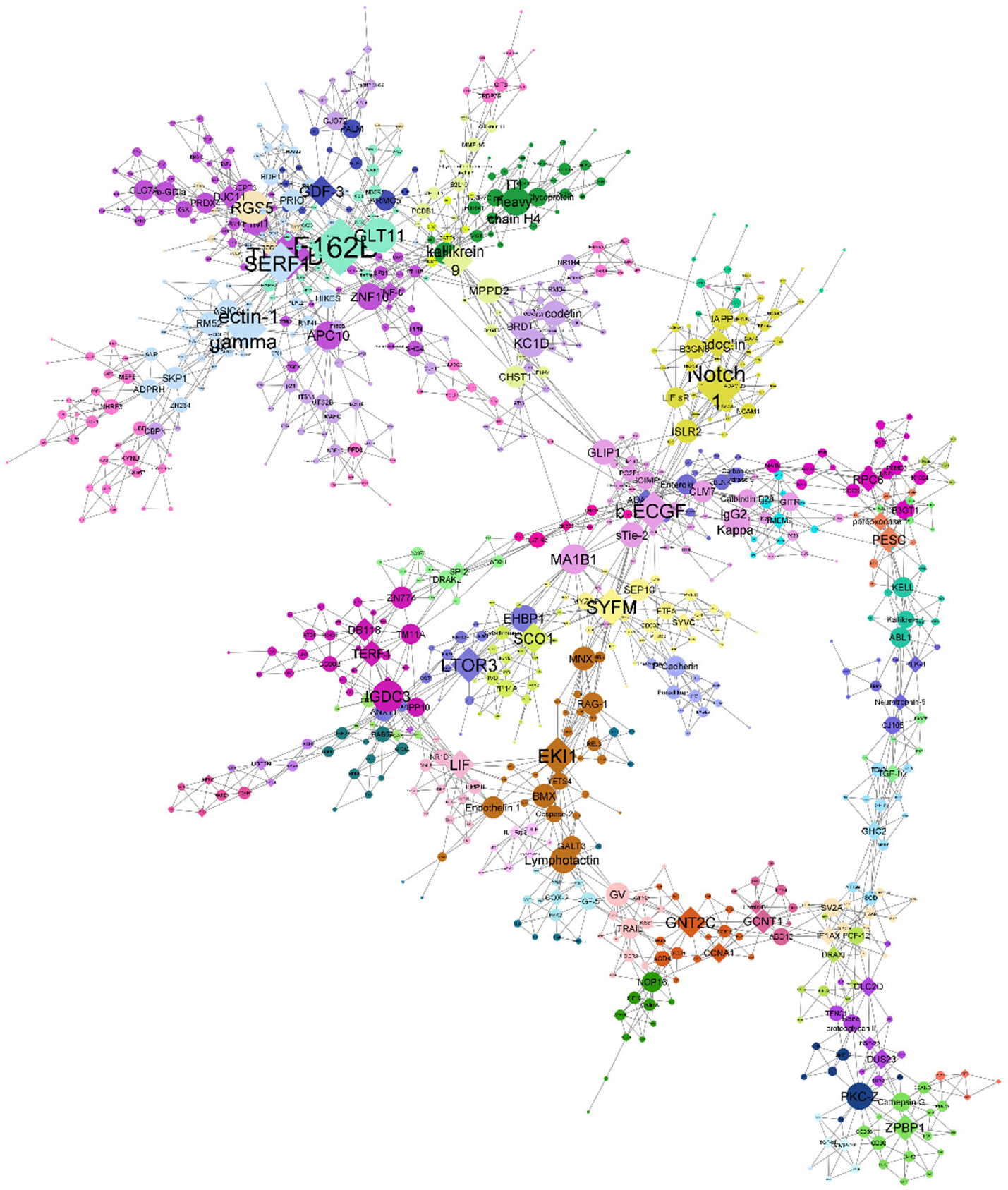
MEGENA network plot of 833 proteins differentially abundant in ventilation status. MEGENA network plot produced using the igraph object produced by MEGENA analysis of 833 proteins differentially expressed in COVID-19-infected individuals who were placed on a ventilator vs those who were not. Colors represent clusters of proteins identified by MEGENA. Shape corresponds to hub protein status; diamonds represent hubs, all other proteins are circular. Node size corresponds to the connectedness of that protein as determined by significant correlations, scaled by number of edges connecting that node.

**Fig. S13.**
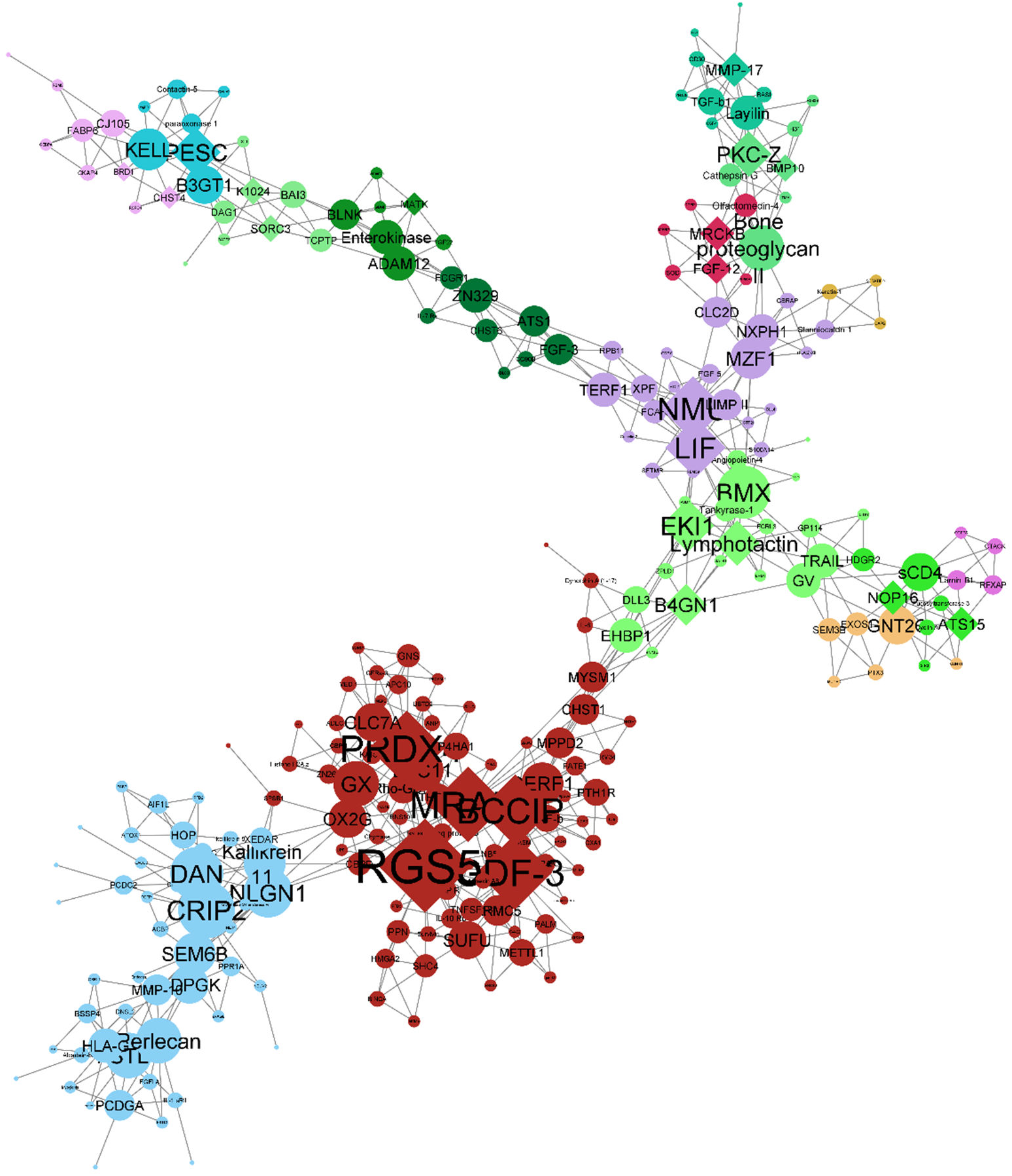
MEGENA network plot of 253 proteins differentially abundant in death status. MEGENA network plot produced using the igraph object produced by MEGENA analysis of proteins differentially expressed in any of case vs control status, ventilation status, or death status. Colors represent clusters of proteins identified by MEGENA. Shape corresponds to hub protein status; diamonds represent hubs, all other proteins are circular. Node size corresponds to the connectedness of that protein as determined by significant correlations, scaled by number of edges connecting that node.

