## Supplemental Figures for "Plasma proteomics of SARS-CoV-2 infection and severity reveals impact on Alzheimer and coronary disease pathways"

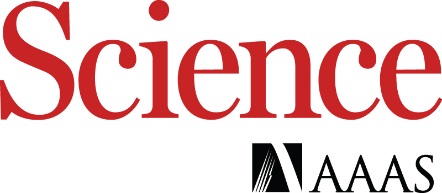

Supplementary Materials for

Plasma proteomics of SARS-CoV-2 infection and severity reveals impact on Alzheimer and coronary disease pathways

Lihua Wang^1,2*^, Dan Western^1,^^2*^, Jigyasha Timsina^1,2^, Charlie Repaci^1,2,10^, Won-Min Song^3^, Joanne Norton^1,2^, Pat Kohlfeld ^1,2^, John Budde^1,2^, Sharlee Climer^4^, Omar H. Bbutt^5^, Daniel Jacobson^6^, Michael Garvin^6^, Alan R Templeton^7^, Shawn Campagna^8^, Jane O’Halloran^9^, Rachel Presti^9^, Charles W. Goss^10^, Philip A. Mudd^11^, Beau M. Ances^5^, Bin Zhang^3^, Yun Ju Sung^1,2,10¶^, Carlos Cruchaga^1,2,12¶^

**This PDF file includes:**

Figs. S1 to S13

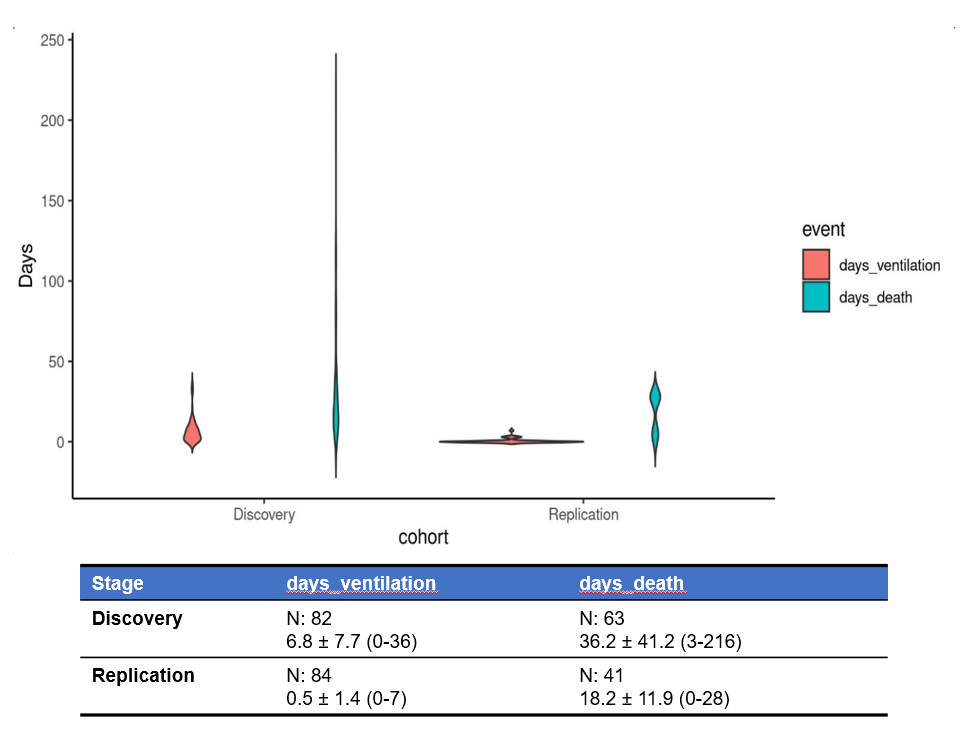

Fig. S1. Violin plots of number of days to ventilation and death in discovery and replication. The table of sample size (N) and mean and standard deviation (mean ± SD) for number of days to ventilation and number of days to death.

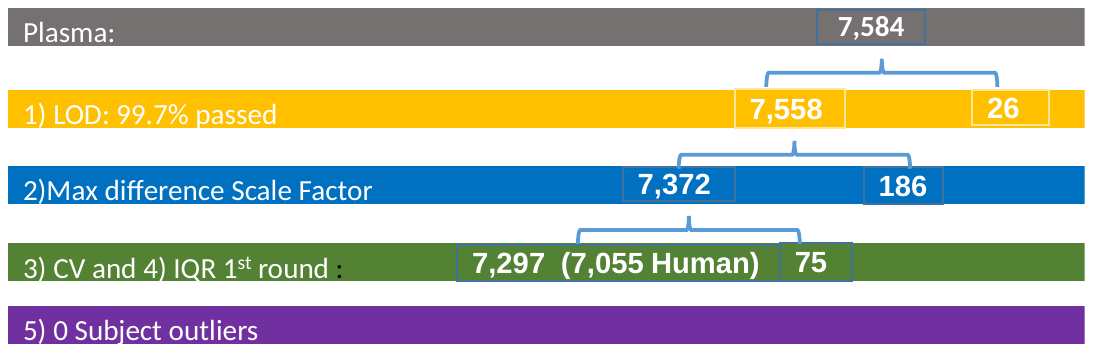

Fig. S2. Quality control flow for plasma SomaScan v4.1 7K data in discovery stage. The number of aptamer kept (left side number) and removed (right side number) in each step was include in this figure. In summary, 7,297 (92.8%) aptamers were kept. Among the 7,297 aptamers, 7,055 which targets human proteins were used in our analyses and 4,301 were available in replication data. LOD represents limit of detection. CV denotes coefficient of variation. The detailed explanation of quality control for each step can be found in Materials and Methods.

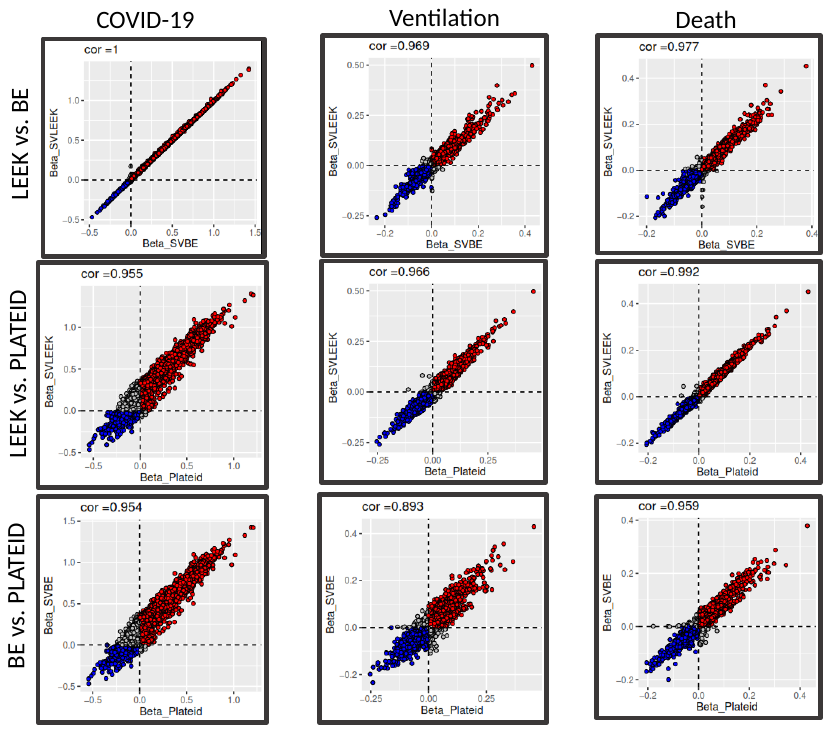

Fig. S3. Scatterplot of effect size for including 3 “LEEK” method estimated surrogate variables, 29 “BE” method estimated surrogate variables and Plateid as covariates in the model. The surrogate variables were estimated using 7,055 proteins assayed for 482 samples using R package sva. Then linear regression was performed including 3 “LEEK” method estimated surrogate variables, or 29 “BE” method estimated surrogate variables, or plate (PLATEID) as covariates to estimate the effect size for 3 outcomes. The effect size from the 3 approaches were similar with very high Pearson correlation coefficient among them.

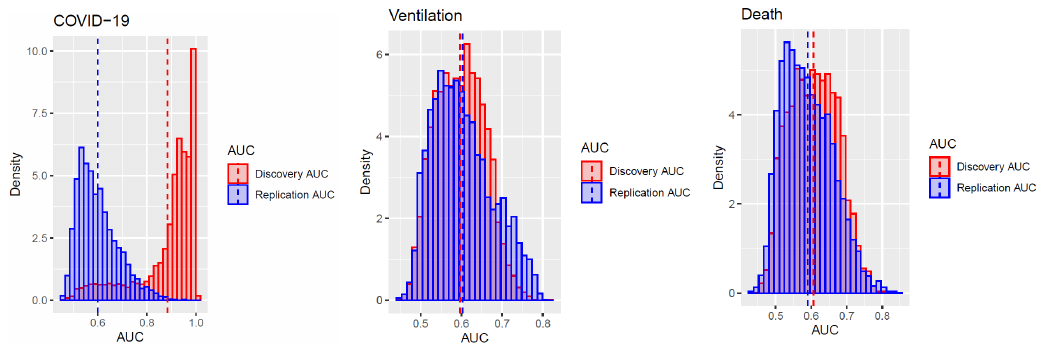

Fig. S4. Distribution of AUC for COVID-19 infection, Ventilation status and Death status. AUC was estimated separately by discovery and replication stage from univariate logistic regression model using each of COVID-19 infection, Ventilation status and Death status as the response variable, and each of 4301 proteins as the predictor. The histograms of AUC (AUC displayed along x-axis and Density displayed along y-axis) were plotted separately for COVID-19 infection, Ventilation status and Death status using ggplot2 R package (red color: AUC in discovery stage; blue color: AUC in replication stage).

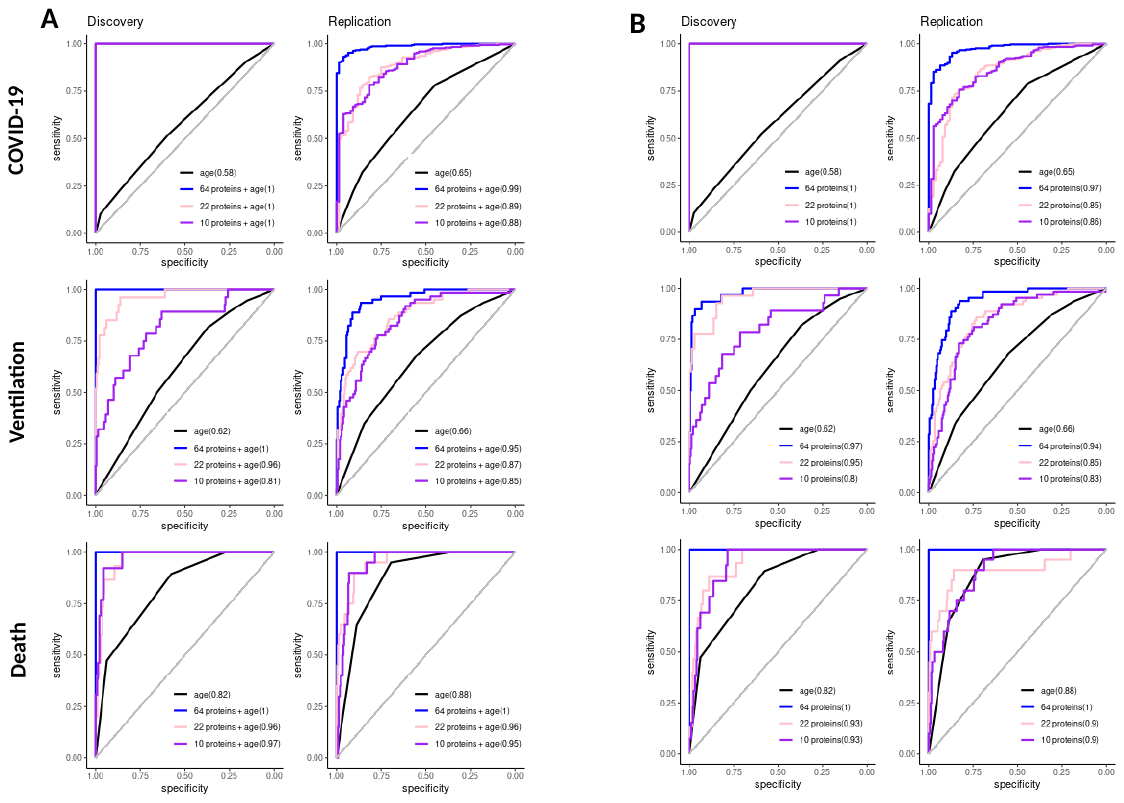

Fig. S5. The ROC curves using 64 significant proteins consistent across all 3 outcomes, 22 proteins selected by Lasso regression for ventilation, and 10 proteins selected by Lasso regression for death. A) The ROC curves with corresponding AUC in each of discovery and replication stage derived from logistic regression model (univariate or multivariate) using each of COVID-19, ventilation and death as response variables, either age along (black solid curves) as predictor, or age together with 64 ventilation proteins (blue solid curves) as predictors, or  age together with 22 proteins (pink solid curves) as predictors, or age together with 10 proteins (purple solid curves) as predictors. Y-axis represents sensitivity and x-axis represents specificity. All these 3 sets of proteins demonstrated significantly higher AUCs (0.81-1) for ventilation and death than the model using age alone.  B) The ROC curves with corresponding AUC in each of discovery and replication stage derived from logistic regression model (univariate or multivariate) using each of COVID-19, ventilation and death as response variables, either age along (black solid curves) as predictor, or 64 ventilation proteins (blue solid curves) as predictors, or  22 proteins (pink solid curves) as predictors, or 10 proteins (purple solid curves) as predictors. Y-axis represents sensitivity and x-axis represents specificity. All these 3 protein set also demonstrated significantly higher AUCs (0.8-1) for ventilation and death than the model using age.

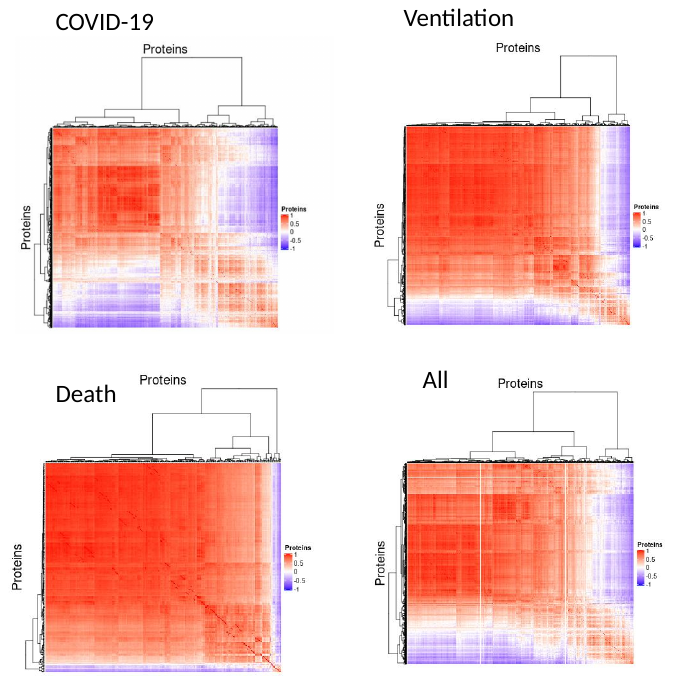

Fig. S6. Heatmap of Pearson correlation matrix of detected and replicated significant proteins for COVID-19, Ventilation, Death and all 3 outcomes combined. Pearson correlation matrix for the significant differential abundant proteins (841 for COVID-19, 831 for ventilation, 253 for death and 1449 for all 3 outcomes combined). ComplexHeatmap R package was used to plot the heatmap.

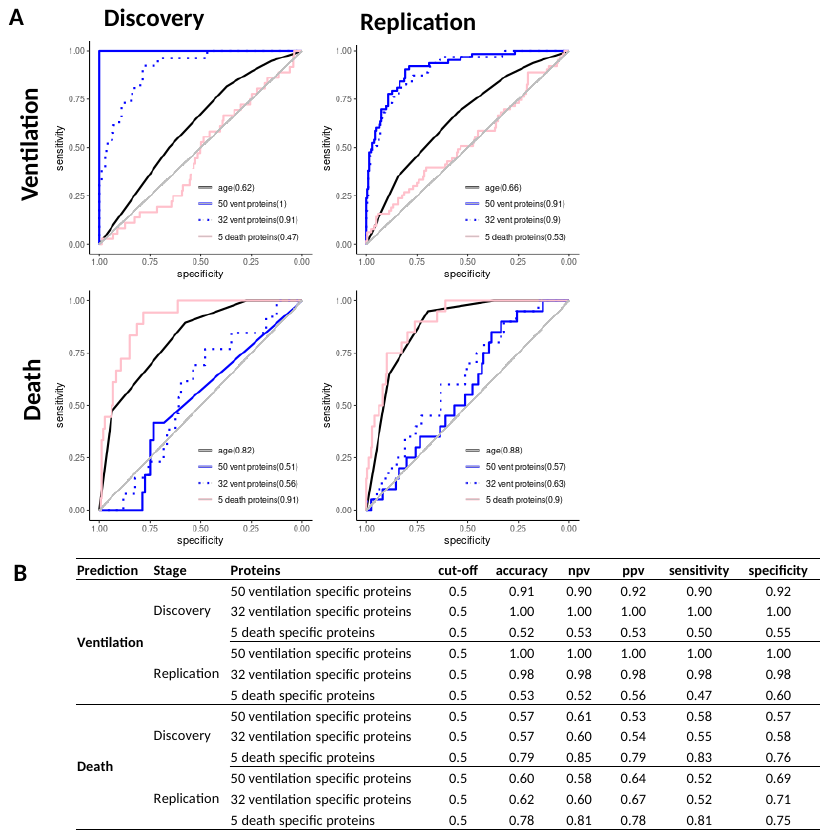

Fig. S7. Performance of prediction model using ventilation specific and death specific proteins. A) The ROC curves with corresponding AUC in each of discovery and replication stage derived from logistic regression model (univariate or multivariate) using each of ventilation and death as response variables, either age along (black solid curves) as  predictor, or 50 ventilation specific proteins (blue solid curves) as predictors, or  32 Lasso selected ventilation specific proteins (blue dotted curves) as predictors, or 5 death specific proteins (pink solid curves) as predictors. Y-axis represents sensitivity and x-axis represents specificity. B) The table of accuracy, negative predictive value.(NPV), positive prediction value (PPV), sensitivity and specificity at 0.5 of Youden's J statistic for the logistic regression model used in b. This evaluation used the case control balanced subsample by selecting age and gender matched or age matched controls in discovery and replication respectively for each of ventilated cases or died cases.  As expected, for ventilation prediction, both 50 and 32 ventilation specific proteins demonstrated ~90% -100% of these evaluations. Whereas for death, 5 death specific proteins showed ~75%-85% of these evaluations.

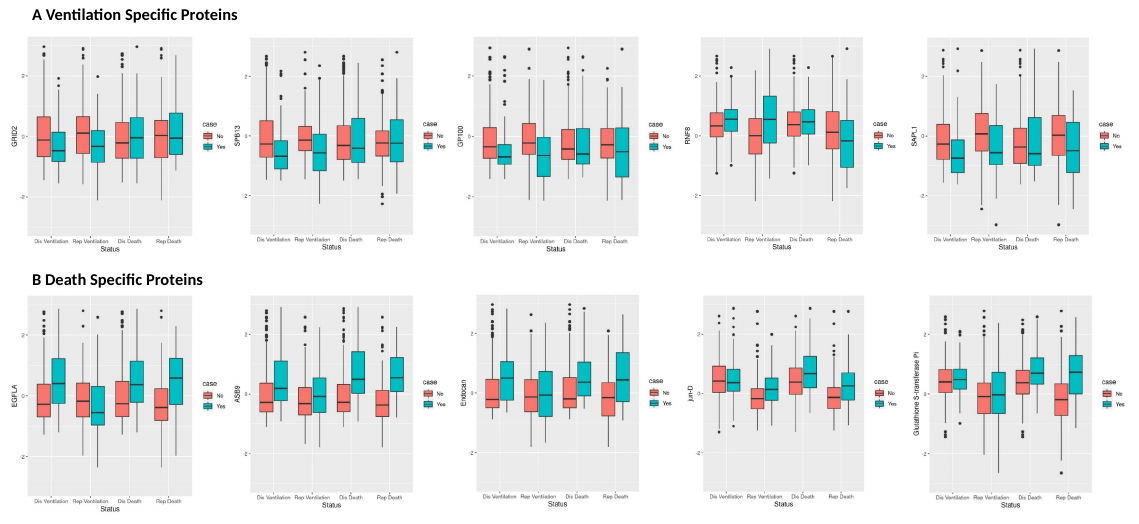

Fig. S8. Boxplots of Ventilation specific proteins and Death specific proteins. A) Boxplots of 5 ventilation specific proteins (GRID2, SPB13, GP100, RNF8, SAPL1) by Status (Dis Ventilation denotes Ventilation status in Discovery; Rep Ventilation represents Ventilation status in Replication; Dis Death denotes Death status in Discovery; Rep Death represents Death status in replication). The y-axis was Z-Score of protein levels which converted separately in discovery stage and replication stage. Horizontal bars demonstrate the median value. The lower and upper borders of the box represent the first and the third quantitle. Dis: Discovery; Rep: Replication. B) Boxplots of 5 death specific proteins (EGFLA, ASB9, Endocan, jun-D, Glutathione S-transferase Pi) by Status (Dis Ventilation denotes Ventilation status in Discovery; Rep Ventilation represents Ventilation status in Replication; Dis Death denotes Death status in Discovery; Rep Death represents Death status in replication). The y-axis was Z-Score of protein levels which converted separately in discovery stage and replication stage. Horizontal bars demonstrate the median value. The lower and upper borders of the box represent the first and the third quantitle. Dis: Discovery; Rep: Replication

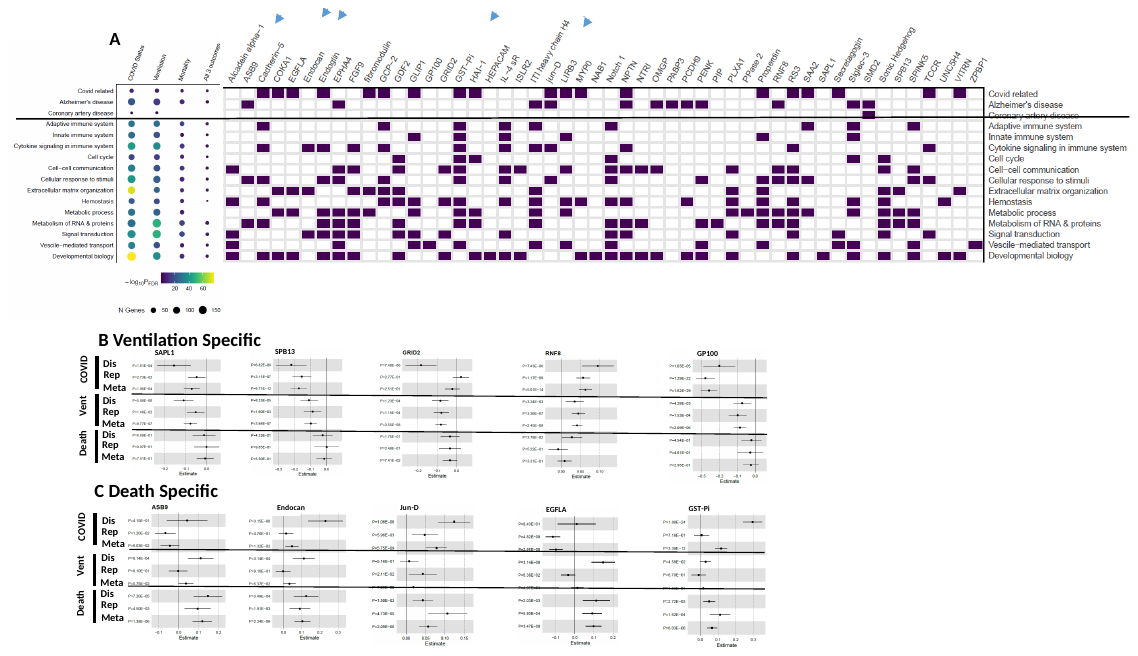

Fig. S9. Pathway enrichment analyses using the Entrez Gene Symbol of the identified significant proteins (45 ventilation specific proteins and 5 death specific proteins included in the figure). A) Entrez gene symbol of the identified significant proteins were used in pathway enrichment analyses via Enrichr and FUMA. 45 ventilation specific proteins and 5 death specific proteins were enriched for 16 pathways. 19 of these proteins were enriched in covid related pathways, 12 of these proteins were enriched in Alzheimer’s disease. And one protein SMD2 (*SNRPD2*) was involved in Coronary artery disease. Blue arrows point to the death specific proteins. B) Forest plots of ventilation specific proteins: Proactivator polypeptide-like 1 (SAPL1), Serpin B13 (SPB13), Glutamate receptor ionotropic, delta-2 (GRID2), E3 ubiquitin-protein ligase RNF8 (RNF8), Melanocyte protein PMEL (GP100) C) Forest plots of death specific proteins: Ankyrin repeat and SOCS box protein 9 (ASB9), Endothelial cell-specific molecule 1 (Endocan), Transcription factor JunD (jun-D), Pikachurin (EGFLA), Glutathione S-transferase Pi (GST-Pi).

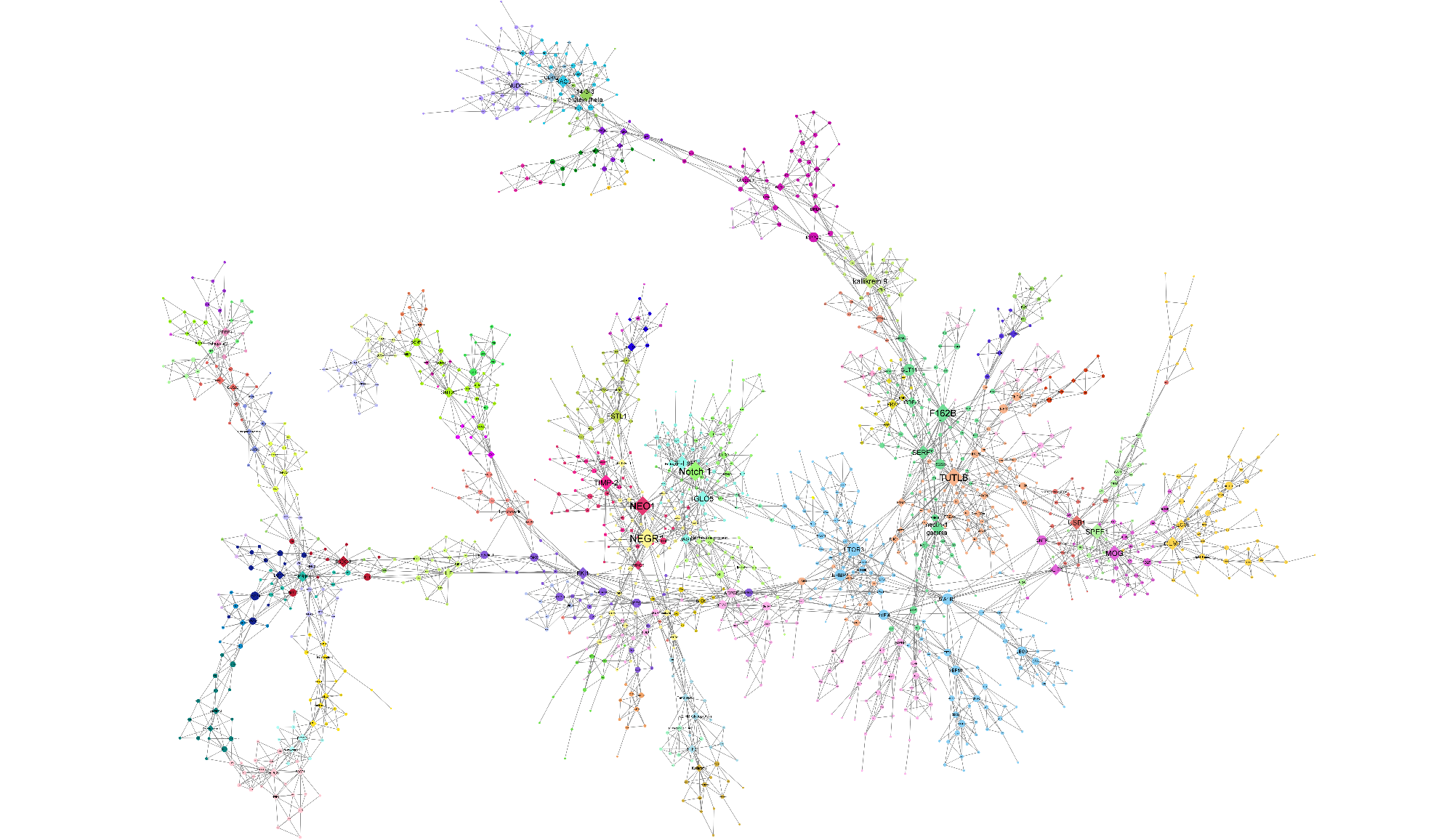

Fig. S10. MEGENA network plot of 1,449 differentially abundant proteins in any of three analyses. MEGENA network plot produced using the igraph object produced by MEGENA analysis of proteins differentially expressed in any of case vs control status, ventilation status, or death status. Colors represent clusters of proteins identified by MEGENA. Shape corresponds to hub protein status; diamonds represent hubs, all other proteins are circular. Node size corresponds to the connectedness of that protein as determined by significant correlations, scaled by number of edges connecting that node.

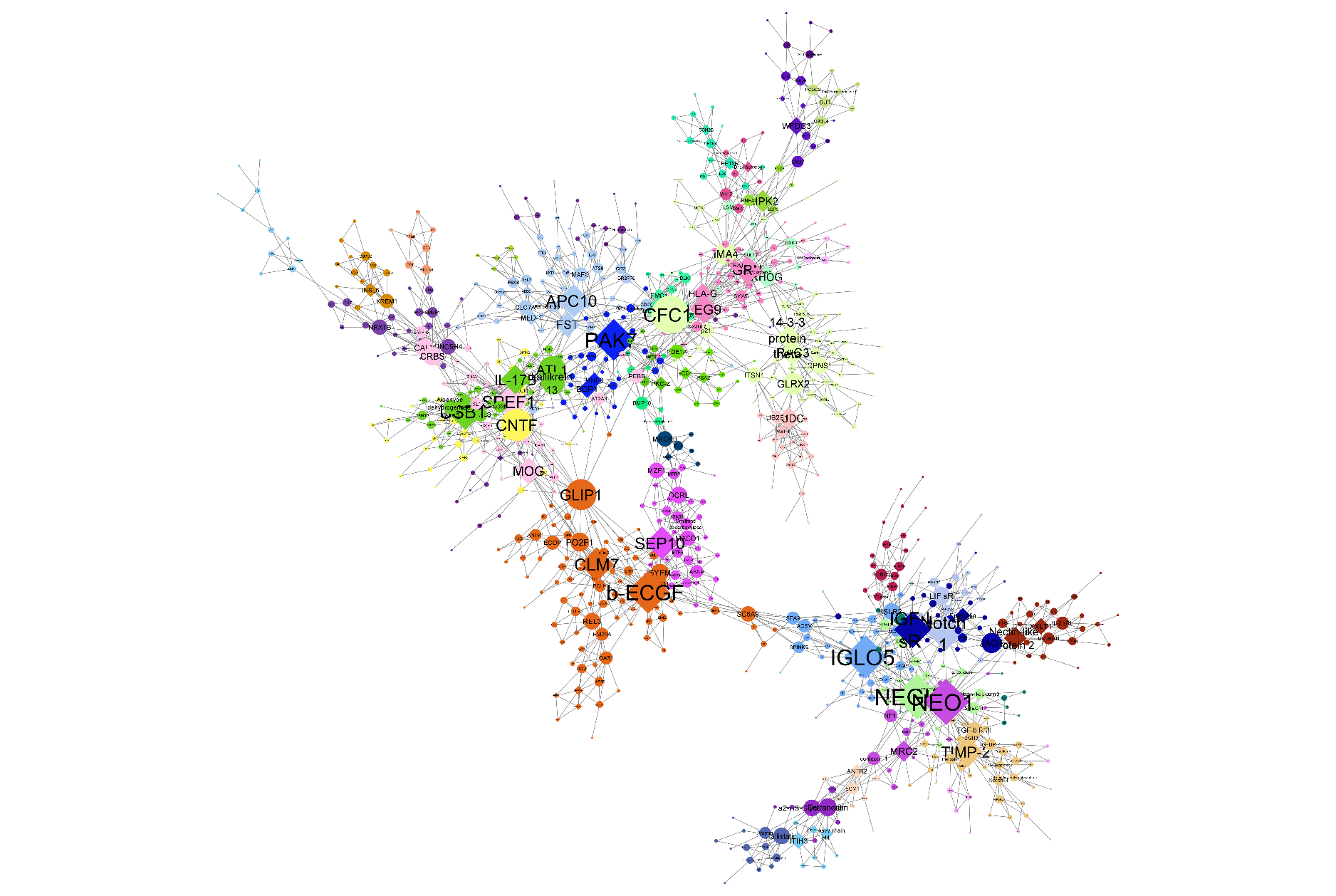

Fig. S11. MEGENA network plot of 841 proteins differentially abundant in COVID-19 case vs control status. MEGENA network plot produced using the igraph object produced by MEGENA analysis of 841 proteins differentially expressed in COVID-19-positive individuals vs unaffected controls. Colors represent clusters of proteins identified by MEGENA. Shape corresponds to hub protein status; diamonds represent hubs, all other proteins are circular. Node size corresponds to the connectedness of that protein as determined by significant correlations, scaled by number of edges connecting that node.

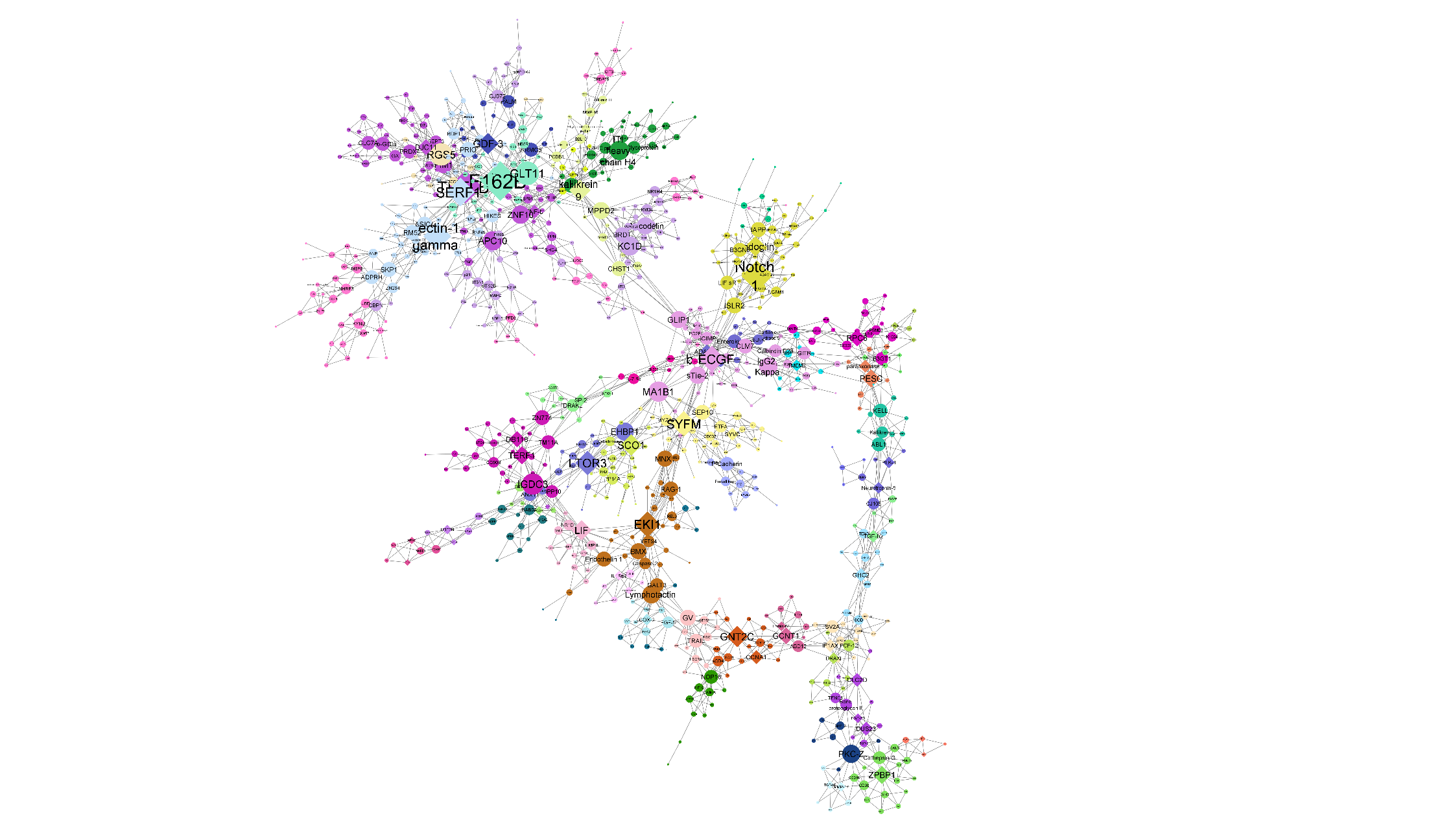

Fig. S12. MEGENA network plot of 833 proteins differentially abundant in ventilation status. MEGENA network plot produced using the igraph object produced by MEGENA analysis of 833 proteins differentially expressed in COVID-19-infected individuals who were placed on a ventilator vs those who were not. Colors represent clusters of proteins identified by MEGENA. Shape corresponds to hub protein status; diamonds represent hubs, all other proteins are circular. Node size corresponds to the connectedness of that protein as determined by significant correlations, scaled by number of edges connecting that node.

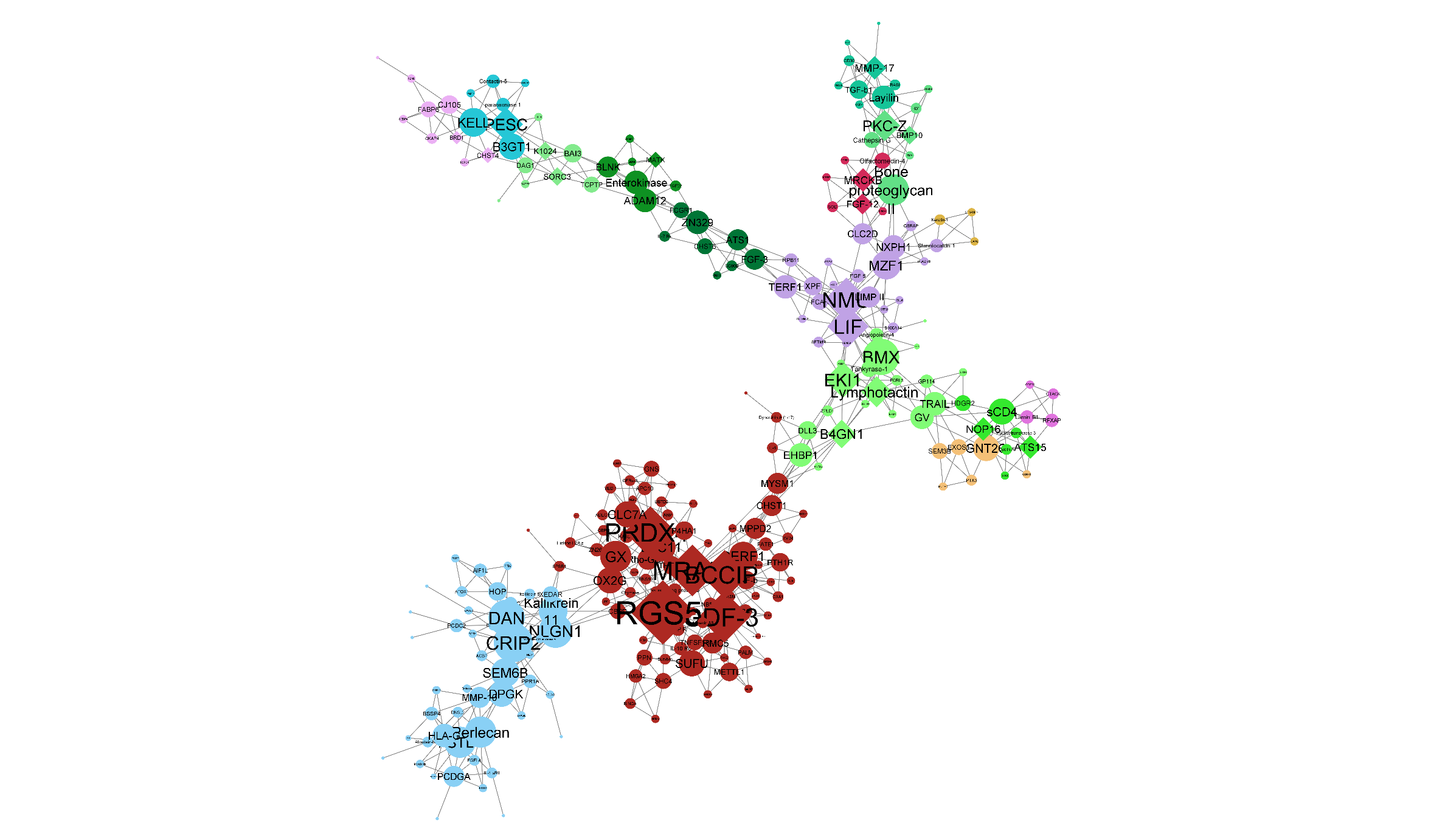

Fig. S13. MEGENA network plot of 253 proteins differentially abundant in death status. MEGENA network plot produced using the igraph object produced by MEGENA analysis of proteins differentially expressed in any of case vs control status, ventilation status, or death status. Colors represent clusters of proteins identified by MEGENA. Shape corresponds to hub protein status; diamonds represent hubs, all other proteins are circular. Node size corresponds to the connectedness of that protein as determined by significant correlations, scaled by number of edges connecting that node.
